# Applications of Fairness Training in Artificial Intelligence to Ophthalmic Disease Diagnosis using External Eye Photographs

**DOI:** 10.64898/2026.11.08.26363489

**Authors:** Vasil Kostin, Sung-Hoon Yoon, Jimmy S. Chen, Seungjoo Baek, Jesse Seid, Nathan Ouyang, Edward Li, Eric Li, Min Shi, Mengyu Wang

**Author notes:** These three authors contributed equally and should be considered co-first authors (Vasil Kostin, Sung-Hoon Yoon, Jimmy S. Chen). Corresponding author: Mengyu Wang, PhD, Harvard Ophthalmology AI Lab, Schepens Eye Research Institute of Massachusetts Eye and Ear, Harvard Medical School, Boston, MA, USA.

## Abstract

Diabetic retinopathy (DR), age-related macular degeneration (AMD), and glaucoma are leading causes of preventable blindness, yet whether external eye photographs carry phenotypic information about posterior segment disease remains unclear, and methods to mitigate demographic disparities in such models are largely unexplored. We developed and evaluated deep learning and vision-language models for binary classification of vision-threatening DR (VTDR), advanced AMD, and glaucoma using 40,687 external eye photographs from 9,878 patients at Massachusetts Eye and Ear. Convolutional networks (WResNet, EfficientNet), a vision transformer (ViT), and a finetuned vision-language model (LLaVA) were assessed by area under the receiver operating characteristic curve (AUC), with fairness across sex, race, and ethnicity quantified by the equity-scaled AUC (ES-AUC). Five fairness-training strategies were applied to the best models. LLaVA achieved the highest baseline AUC for VTDR (85·4%, comparable to ViT 82·6%); advanced AMD was similar across architectures (EfficientNet 86·3%; LLaVA 84·8%; ViT 84·7%), whereas glaucoma was substantially worse (67-74%). Fairness training improved VTDR and AMD AUCs to 86·3% and 85·9% while partially mitigating demographic disparities. In summary, although AI models that rely on external eye photography to detect posterior segment disease may not yet be suitable for clinical deployment, the fairness methods demonstrated in this proof-of-concept study could be applied to other AI systems to decrease performance disparities and potentially improve model performance.

## Introduction

Diabetic retinopathy (DR), age-related macular degeneration (AMD), and glaucoma are among the leading causes of blindness in the United States, collectively affecting over 20 million Americans.[1] Due to the high volume nature of ophthalmology and widespread use of in-clinic imaging, artificial intelligence (AI) is poised to augment ophthalmic clinical care, including disease diagnosis and predicting disease progression and forecasting. Specifically, there exist several high-performing models for diagnosis of posterior segment disease such as DR, glaucoma, and AMD using fundus photographs, optical coherence tomography (OCT), and more recently optical coherence tomography angiography (OCTA). More recently, there has been growing interest in whether deep learning can identify latent disease-associated information or systemic data from imaging modalities not traditionally used for standard-of-care diagnosis.[2, 3]

External eye photographs represent a unique opportunity for AI-based analysis of periocular and orbital biomarkers associated with intraocular disease and systemic health status. Prior work using external eye photographs has focused largely on identifying eyelid measurements and oculo-facial disease[4, 5], strabismus screening[6], and various systemic laboratory abnormalities and diabetic retinopathy.[2, 3] Deep learning applied to external eye photographs can estimate circulating biomarkers such as glycated haemoglobin (HbA1c) and blood lipids.[3, 2] Elevated HbA1c is an independent risk factor for vision-threatening diabetic retinopathy,[7] and systemic changes in lipid metabolism have been linked to AMD, to disease stage, and to specific retinal imaging features.[8, 9, 10] We therefore hypothesised that external eye photographs may carry phenotypic signal associated with posterior-segment disease, including vision-threatening DR and advanced AMD, reflecting shared systemic and periocular correlates, even though these posterior structures are not directly imaged. Recent studies have also applied deep learning to anterior segment OCT and corneal topography to predict demographic characteristics such as age and sex.[11, 12] Other studies have explored external eye features associated with systemic biomarkers, including HbA1c and haemoglobin,[13] and conjunctival vascular changes in diabetes.[14]

A well-known issue with fine-tuning AI models for diagnostic prediction is variable performance across important dataset characteristics, such as race and sex. This is particularly concerning in healthcare because DL models that perform variably across populations, especially underrepresented groups, have the potential to propagate disparities.[15] For example, prior work has demonstrated that DL models tend to misclassify skin diseases such as melanoma in patients with darker skin tones.[16] Similar biases have been exposed in DL models predicting heart failure hospitalisation length[17] and postpartum depression.[18] To address these concerns, methods to mitigate bias beyond simple dataset augmentation have been explored to optimise models more fairly across demographic groups, collectively termed “fairness training”.[19]

There currently are few proof-of-concept studies implementing fairness training for DL models in ophthalmology.[20, 21] These studies evaluated models on fundus photographs using fairness training to fine-tune foundation models and convolutional neural network architectures. However, there remains a paucity of literature describing how fairness methods could improve DL model performance on external eye photographs. There also exists a gap in knowledge regarding how fairness training could optimise current state-of-the-art models such as vision transformers (ViT)[22] and vision-language models (VLMs).

In this proof-of-concept study, we demonstrate the utility of several state-of-the-art DL models in classifying vision-threatening DR, advanced AMD, and glaucoma using external eye photographs (Figure 1). We compare the performances of WResNet,[23, 24] EfficientNet,[25] ViT,[22] a VLM (LLaVA)[26]. We additionally evaluated several fairness training methods on demographic features including race, sex, and ethnicity: distributionally robust optimisation (DRO),[27] adversarial learning (Adv),[28] fair supervised contrastive loss (FSCL),[29] fair identity scaling (FIS),[30] and FairLoRA for VLMs [31]. We assessed disease-specific performance using the area under the receiver operating characteristic curve (AUC) and sensitivity at various thresholds, as well as demographic-specific performance using the equity-scaled AUC (ES-AUC).[21]

**Figure 1:**
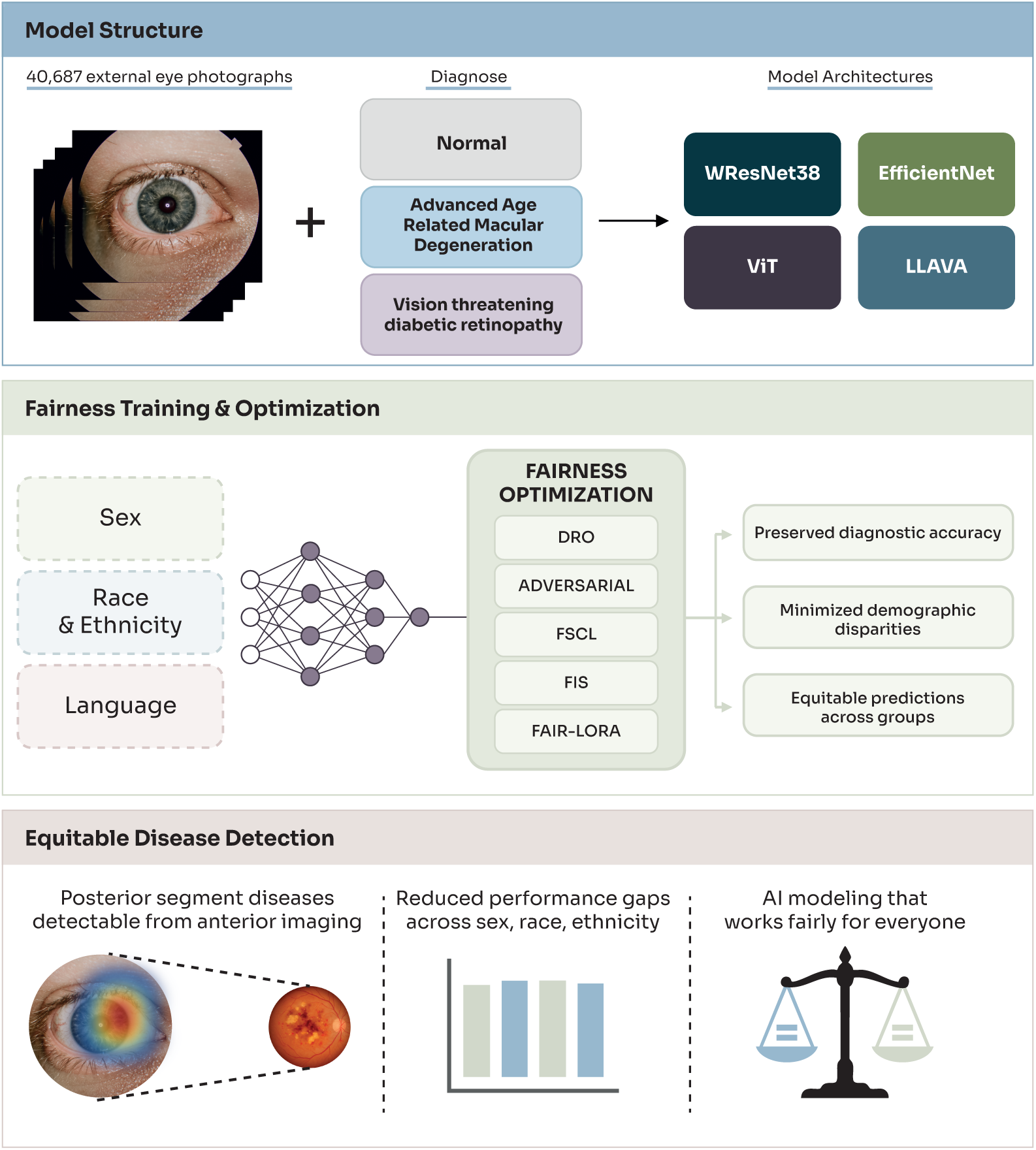
Study overview. External eye photographs from 40,687 images were labeled using ICD codes for vision-threatening diabetic retinopathy, advanced age-related macular degeneration, and normal status. Four model architectures (WResNet38, EfficientNet, ViT, and LLaVA) were trained and evaluated. Fairness optimisation methods (DRO, adversarial debiasing, FSCL, FIS, FairLoRA) were applied to reduce performance disparities across sex, race, and ethnicity, achieving equitable disease detection from anterior segment imaging.

## Results

### Study Population

We analysed 40,687 external eye photographs from 9,878 unique patients. The demographic characteristics are summarised in Table 1. Among all participants, 57·2% (n=5,647) were female and 42·8% (n=4,230) were male. The racial composition included 80·1% White (n=7,916), 12·4% Other/Unknown (n=1,223), 4·1% Black (n=407), and 3·4% Asian (n=332). Regarding ethnicity, 88·8% (n=8,768) identified as non-Hispanic, 5·6% (n=556) as Hispanic, and 5·6% (n=554) had unknown ethnicity.

**Table 1:** Demographic and clinical characteristics of the study population. Data are presented as n (%). The dataset was split into training and test sets at approximately 1:1 ratio. Disease prevalence is reported at the patient level.

| **Characteristic** | **Training Set** | **Test Set** | **Total** |
|---|---|---|---|
| **Photographs, n** | 20,307 | 20,380 | 40,687 |
| **Unique patients, n** | 4,939 | 4,939 | 9,878 |
| **Sex** |  |  |  |
| Female | 2,843 (57.6%) | 2,804 (56.8%) | 5,647 (57.2%) |
| Male | 2,095 (42.4%) | 2,135 (43.2%) | 4,230 (42.8%) |
| **Race** |  |  |  |
| White | 3,963 (80.2%) | 3,953 (80.0%) | 7,916 (80.1%) |
| Black | 199 (4.0%) | 208 (4.2%) | 407 (4.1%) |
| Asian | 166 (3.4%) | 166 (3.4%) | 332 (3.4%) |
| Other/Unknown | 611 (12.4%) | 612 (12.4%) | 1,223 (12.4%) |
| **Ethnicity** |  |  |  |
| Non-Hispanic | 4,370 (88.5%) | 4,398 (89.0%) | 8,768 (88.8%) |
| Hispanic | 291 (5.9%) | 265 (5.4%) | 556 (5.6%) |
| Unknown | 278 (5.6%) | 276 (5.6%) | 554 (5.6%) |
| **Age at first image, years**<sup>*b*</sup> |  |  |  |
| Mean (SD) | – | – | 59.7 (18.8) |
| Median (IQR) | – | – | 62.9 (48.9–73.1) |
| <18 | 155 (3.1%) | 151 (3.1%) | 306 (3.1%) |
| 18–39 | 634 (12.8%) | 649 (13.1%) | 1,283 (13.0%) |
| 40–64 | 1,890 (38.3%) | 1,905 (38.6%) | 3,795 (38.4%) |
| 65–79 | 1,664 (33.7%) | 1,618 (32.8%) | 3,282 (33.2%) |
| ≥80 | 596 (12.1%) | 616 (12.5%) | 1,212 (12.3%) |
| **Disease prevalence (patients)** |  |  |  |
| Vision-threatening DR | 50 (1.0%) | 65 (1.3%) | 115 (1.2%) |
| Advanced AMD | 176 (3.6%) | 151 (3.1%) | 327 (3.3%) |
| Glaucoma<sup>*a*</sup> | 366 (8.9%) | 372 (9.0%) | 738 (8.9%) |

<sup>*a*</sup>Glaucoma analysis included 8,261 patients (4,111 training; 4,150 test) with available diagnostic data.
<sup>*b*</sup>Mean, SD, median and IQR are reported for the full cohort; split-specific age bands use age at the patient's first image within that split. No ages were missing. Of the 306 patients younger than 18 years, none was VTDR- or advanced AMD-positive and 17 were glaucoma-positive (Supplementary Table S13).

Disease prevalence at the patient level was as follows: vision-threatening DR 1·2% (n=115 patients), advanced AMD 3·3% (n=327 patients), and glaucoma 8·9% (n=738 patients). Glaucoma analysis was limited to 8,261 patients with available visual field data. Mean age at the first study image was 59·7 years (SD 18·8; median 62·9, IQR 48·9-73·1), with no missing ages; 306 patients (3·1%) were younger than 18 years. Patients with positive findings were unevenly distributed across racial groups: vision-threatening diabetic retinopathy (DR) occurred in four out of 332 Asian patients, 22 out of 407 Black patients, 53 out of 7,916 White patients, and 36 out of 1,223 patients of other or unknown race; advanced AMD occurred in nine, four, 287, and 27 patients, respectively (entire cohort; Supplementary Table S11). The number of images in the test set and the number of positive images by demographic subgroup are presented in Supplementary Table S12, and the age distribution by disease status is presented in Supplementary Table S13.

### Model Performance

For DR, LLaVA achieved the highest AUC (85·4%), comparable to ViT (82·6%, 95% CI 79·9-85·1), with WResNet (78·2%, 74·9-81·4) and Efficient-Net (78·7%, 75·3-81·8) lower. For advanced AMD, EfficientNet achieved the highest AUC (86·3%, 84·5-88·1), with ViT (84·7%, 82·8-86·5), LLaVA (84·8%) and WResNet (83·0%, 80·9-85·0) comparable; overlapping 95% confidence intervals indicated no statistically significant superiority of any single architecture. Glaucoma detection was substantially worse across all models (AUC 67-74%), precluding fairness analysis (Supplementary Table S3). All confidence intervals for the conventional models are from 2,000 patient-clustered bootstrap resamples of the test set (Methods); LLaVA estimates are point values.

At clinically relevant specificity thresholds, LLaVA demonstrated the highest sensitivity for DR detection: 76·7% at 80% specificity, 71·5% at 85% specificity, and 61·7% at 90% specificity. For AMD, performance was comparable across architectures with sensitivities of 70-74% at 80% specificity, though LLaVA demonstrated the highest sensitivities (65.1%, 55.1%, 36.4%) at the 85%, 90%, and 95% thresholds respectively (Figure 2).

**Figure 2:**
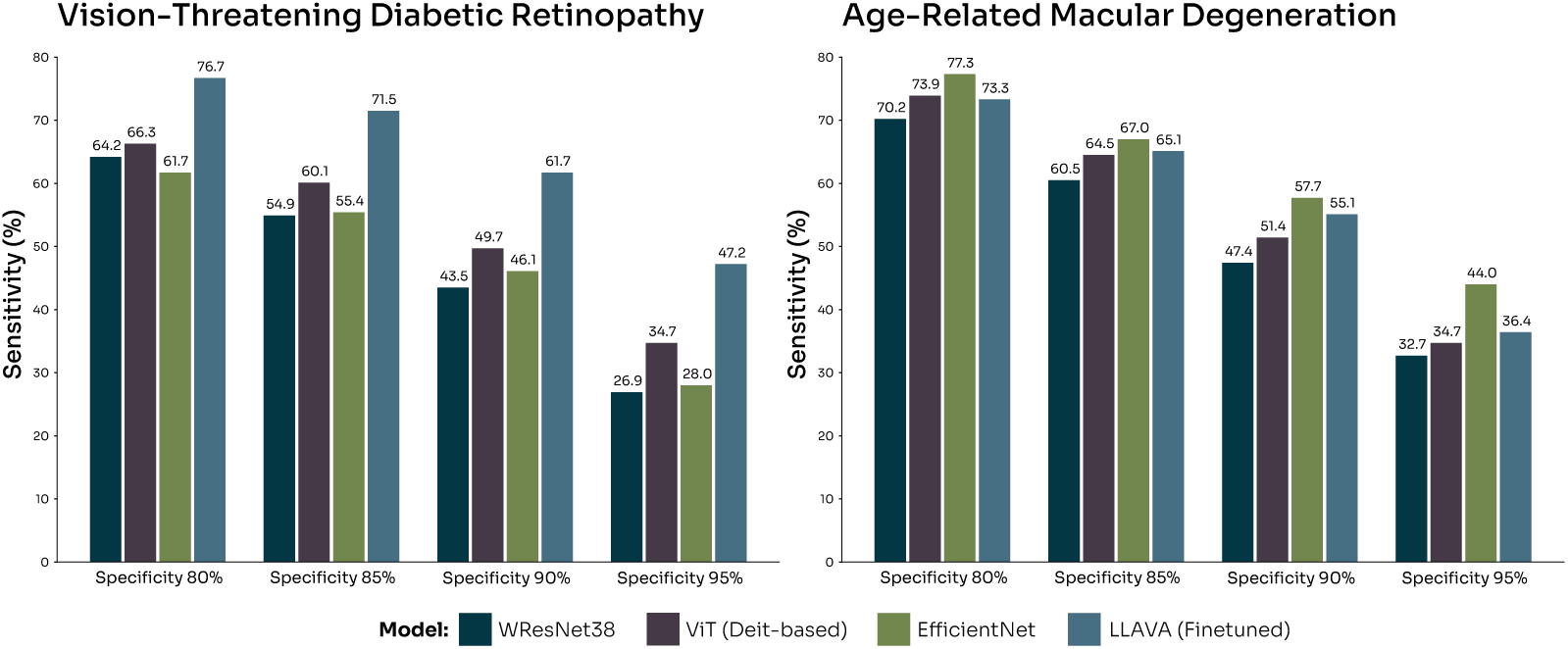
Model sensitivity at fixed specificity thresholds. Sensitivity for detecting vision-threatening diabetic retinopathy (left) and age-related macular degeneration (right) at specificity thresholds of 80%, 85%, 90%, and 95%. WResNet38 = Wide ResNet-38; ViT = Vision Transformer (DeiT-based); LLaVA = Large Language and Vision Assistant.

### Performance in undilated photographs

Device-stratified results are reported in the Supplementary Information (Supplementary Note and Supplementary Tables S14-S16); 98·7% of images were acquired on Topcon-labelled platforms, and within this group the base-line ViT performed as in the full test set (AUC 82·9%, 85·5% and 73·9% for vision-threatening DR, advanced AMD and glaucoma).

Of the 20,380 test images, 19,523 (95·8%) were acquired after pharmaco-logical dilation and 857 (4·2%) were undilated. Evaluated on the undilated subset alone, without retraining, the baseline ViT retained an AUC of 70·8% (95% CI 58·8-82·2; 21 positive images) for vision-threatening DR and 74·0% (65·6-82·2; 26 positive images) for advanced AMD, compared with 83·3% (80·4-85·8) and 85·7% (83·8-87·4) on dilated images; glaucoma performance was unchanged (72·0% versus 74·0%). Several fairness-trained ViT models retained higher undilated performance for DR than the baseline (for example, adversarial debiasing 79·2%, 71·0-86·6, and fair identity scaling 81·3%, 73·3-88·4, for the sex-attribute models). Discrimination therefore persisted, with lower point estimates and wide intervals, in photographs without a dilated pupil, although the undilated subset was small (Supplementary Table S17).

### Demographic Disparities and Pairness Training

Substantial baseline disparities existed, particularly for race. For vision-threatening DR, the ViT baseline had an ES-AUC of 81·4% for sex, 56·9% for race, and 64·3% for ethnicity, the low race value reflecting a large gap between subgroups. In a single training run per method, adversarial de-biasing raised the race ES-AUC from 56·9% (95% CI 49·7-64·8) to 75·6% (64·0-77·0) and reduced the maximum racial disparity from 25·9% to 5·6%; the bootstrap intervals of the baseline and the adversarially trained model overlap only marginally, and the estimate rests on small numbers of positive test images in the minority strata (7 Asian and 45 Black VTDR-positive images; Supplementary Table S12), so we regard this as a large but not conclusively established improvement. Sex equity, already high, changed little (ES-AUC 81·4% to 85·4% with fair identity scaling) and ethnicity changed modestly (64·3% to 67·2%). The overall AUC of the best fairness-trained models reached 85·7% (95% CI 83·1-88·1) for sex and 86·3% for race (Figure 3). For AMD, fairness effects were smaller and mixed: the ViT baseline race ES-AUC was 60·5%, improving modestly to 67·5% with distributionally robust optimisation, whereas sex equity was essentially unchanged (81·8% to 81·9%) and ethnicity did not improve (79·7% at baseline); the race-specific estimates for AMD are based on only two Asian and two Black test images with positive AMD findings and are therefore not interpretable (see Supplementary Table S12) (Figure 4). Complete per-subgroup results for all models, baseline and fairness-trained, are reported in Supplementary Tables S1-S7.

**Figure 3:**
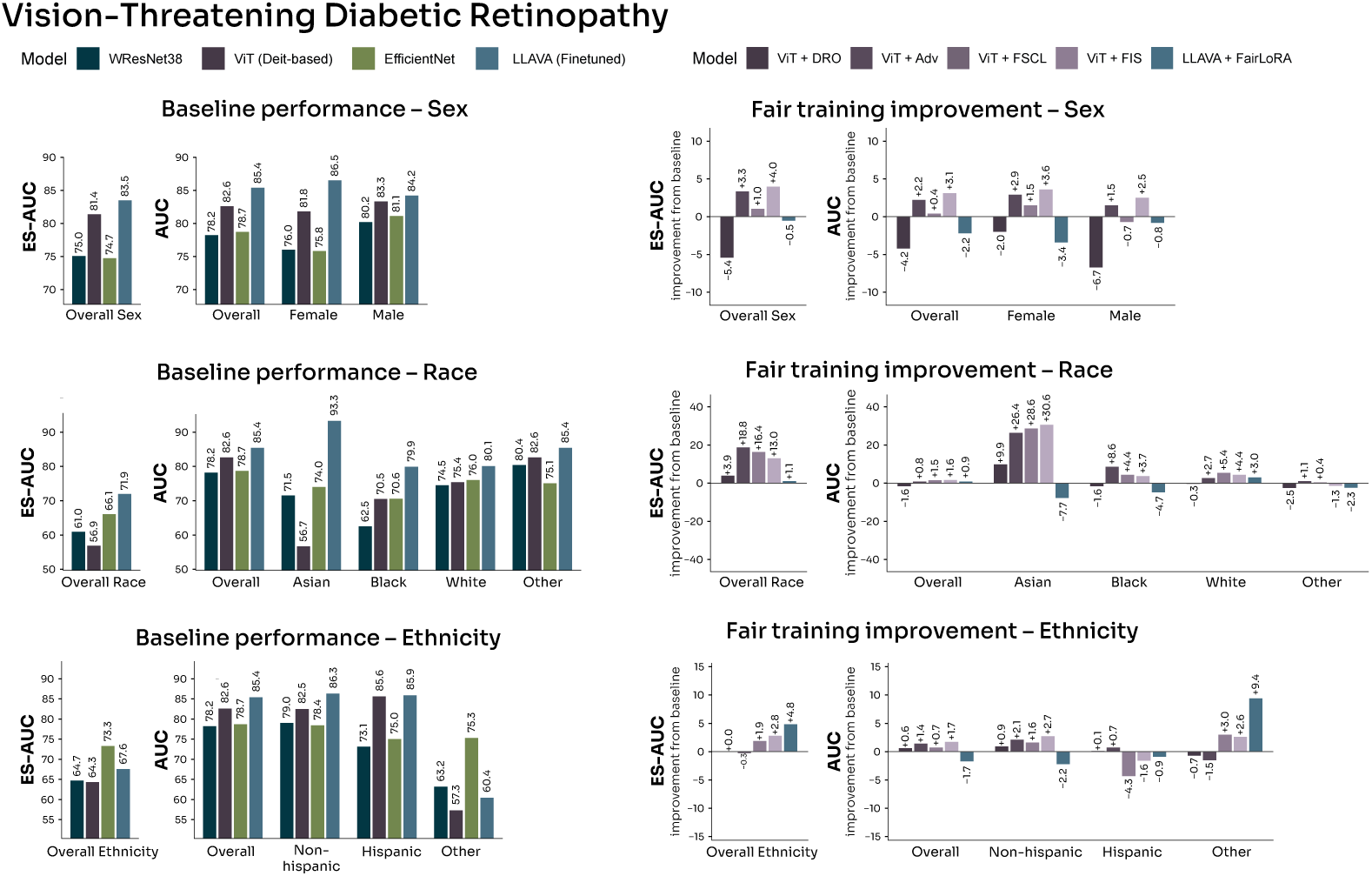
Baseline performance and fairness training improvement for vision-threatening diabetic retinopathy detection. Left panels show baseline model perfor-mance (AUC and ES-AUC) stratified by sex, race, and ethnicity. Right panels show the change in performance after applying fairness training methods. DRO = distributionally robust optimisation; Adv = adversarial debiasing; FSCL = fair supervised contrastive learning; FIS = fair identity scaling.

**Figure 4:**
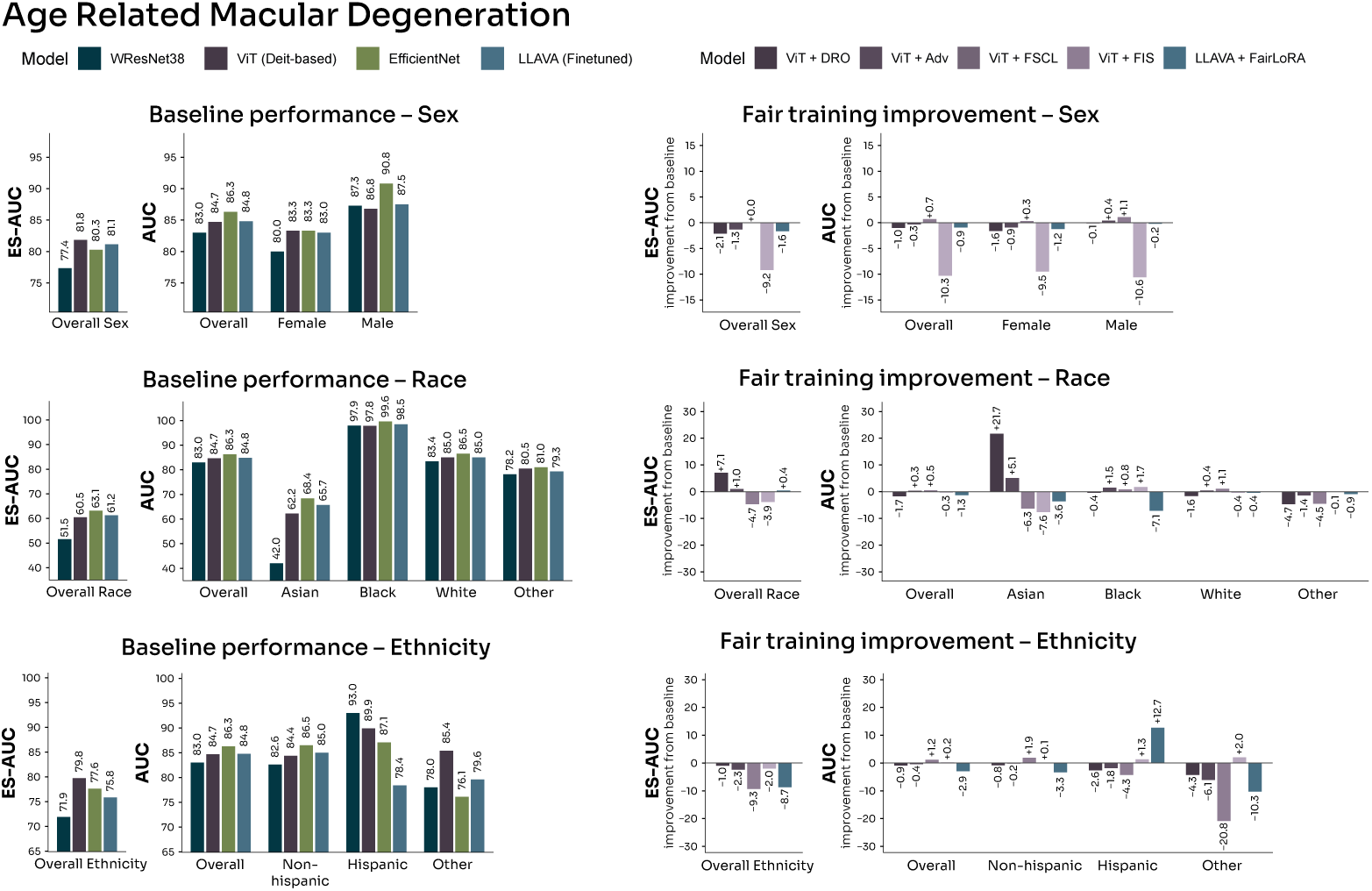
Baseline performance and fairness training improvement for advanced age-related macular degeneration detection. Left panels show baseline model performance stratified by sex, race, and ethnicity. Right panels show the change in performance after applying fairness training methods. DRO = distributionally robust optimisation; Adv = adversarial debiasing; FSCL = fair supervised contrastive learning; FIS = fair identity scaling.

For AMD, fairness training showed smaller improvements. The baseline already had lower ethnic disparity (5·5%), though racial disparity remained high (35·6%) given the small number of minority samples (two Asian and two Black test images with positive AMD findings; see Supplementary Tables S6 and S12).

To distinguish genuine equity gains from generic performance effects, we compared each fairness method against its baseline on overall AUC, equity-scaled AUC, and maximum subgroup disparity (Supplementary Table S10). For DR, adversarial debiasing produced the largest change in racial equity, raising the race ES-AUC from 56·9% (95% CI 49·7-64·8) to 75·6% (64·0-77·0), an increase of 18·7 percentage points, and reducing the maximum racial disparity from 25·9% to 5·6%; fair supervised contrastive learning and fair identity scaling also increased race ES-AUC (to 73·3% [65·0-77·0] and 69·9% [63·0-74·6]) and reduced disparity (to 10·4% and 13·1%), while the accompanying overall-AUC changes were small (e.g., adversarial debiasing +0·8%, 95% CI −1·9 to +2·5; paired comparison with 100 patient-clustered resamples). Because each method was trained once, these comparisons do not account for run-to-run (seed) variability (see Limitations). For AMD, fairness effects were smaller and mixed; several methods altered overall AUC without improving equity, indicating that AMD improvements reflected regularisation more than genuine equity gains.

### Image signal beyond demographic factors

To assess whether model performance reflected genuine image-based signal rather than demographic shortcuts (e.g., inferring age or race from peri-ocular features), we compared each image model against a demographics-only baseline (logistic regression and gradient-boosted trees on age, race, and ethnicity), using the better-performing baseline as the comparator with 100-fold patient-clustered bootstrap 95% confidence intervals and paired tests. For DR, demographics alone achieved an AUC of 74·2%, and all image models significantly exceeded this baseline (ViT +8·4%, 95% CI 5·5-11·8; EfficientNet +4·4%, 0·5-8·1; WResNet +4·0%, 0·4-7·4; all 95% CIs excluding zero), indicating that the DR models captured substantial image-specific information beyond demographic risk. For advanced AMD, demographics alone achieved a higher AUC of 82·6%, reflecting the age dependence of AMD; the image added a smaller but significant increment (EfficientNet +3·8%, 95% CI 2·2-5·6; ViT +2·1%, 0·1-3·8), whereas WResNet did not significantly exceed the baseline (+0·4%, −1·6-2·3). For glaucoma, demographics alone per-formed near chance (AUC 55·0%) and the image models exceeded this base-line by a wide margin (ViT +18·9%, 95% CI 16·3-21·2; EfficientNet +12·7%; WResNet +12·4%; all 95% CIs excluding zero), indicating that external photographs carry glaucoma-associated signal beyond demographics even though absolute performance remained modest (Supplementary Tables S8-S9).

### Class Activation Mapping (OAM)

To understand the features learned by different models, we performed class activation mapping (CAM) analysis for WResNet, EfficientNet, ViT, and LLaVA (Figure 5).

**Figure 5:**
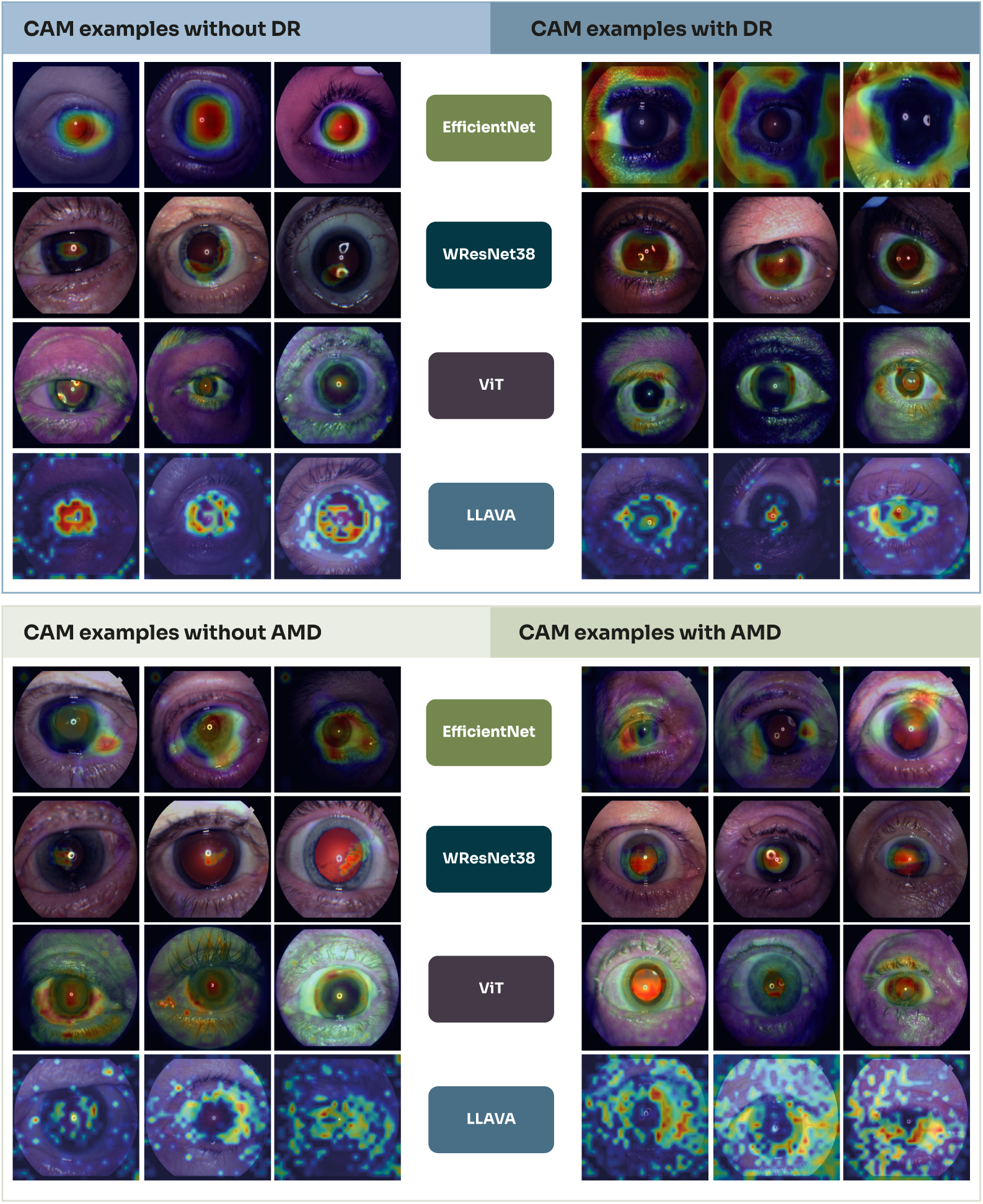
Class Activation Mapping (CAM) visualizations for diabetic retinopathy (DR) and age-related macular degeneration (AMD) detection. Heatmaps illustrate attention patterns for EficientNet, WResNet, ViT, and LLAVA across disease and non-disease cases. Warmer colors (red/yellow) indicate regions of higher model attention.

For vision-threatening DR detection, in non-DR cases EfficientNet focused on the red reflex and the iris-scleral junction without attending to skin or vessels. WResNet similarly attended to the red reflex and iris, with minimal attention to the iris-scleral junction. ViT showed attention primarily toward skin, eyelids, and the red reflex, with some activation on the iris near the iris-scleral junction. LLaVA focused on the red reflex, iris, and iris-scleral junction, with no strong activation from skin regions.

In DR cases, attention patterns shifted notably. EfficientNet redirected attention away from the iris toward the skin, sclera, and occasionally the iris-scleral junction. WResNet maintained attention on the red reflex and iris while extending to vessels and the iris-scleral junction. ViT showed stronger activation toward the iris and sclera, with continued attention to the eyelids. LLaVA concentrated strongly on the iris-scleral junction and the red reflex.

For advanced AMD detection, in non-AMD cases EfficientNet attended to vessels, the red reflex, and diffusely to the iris, with occasional attention to the eyelids. WResNet focused primarily on the red reflex with some attention to the iris. ViT showed the strongest activation toward the iris, sclera, and iris-scleral junction, with additional attention to the eyelids and occasionally the red reflex. LLaVA attended to the iris and iris-scleral junction, with some attention to skin regions.

In AMD cases, EfficientNet shifted attention away from the red reflex to-ward the iris, iris-scleral junction, eyelids, and vessels. WResNet maintained focus on the red reflex with slightly increased attention to the iris compared to non-AMD cases. ViT shifted its strongest attention toward the red reflex, with reduced attention to the iris and occasional attention to the eyelids. LLaVA showed attention toward the iris, sclera, and skin.

## Discussion

In proof-of-concept study, we assessed a spectrum of deep learning model architectures and fairness training methods for detecting vision-threatening DR, advanced AMD, and glaucoma from external eye photographs. Our study has three key findings: (1) DL models demonstrated high performance in identifying vision-threatening DR and AMD from external eye photographs; (2) VLMs such as LLaVA performed comparably to the best conventional architectures, with no statistically significant superiority; and (3) fairness training methods improved both overall performance and subgroup performance for DR across various demographic groups, in single training runs and with the caveats on minority-group case counts discussed below.

Our first key finding was that DL models demonstrated high performance in identifying vision-threatening DR and AMD from external eye photographs. These findings underscore the potential for AI-assisted biomarker exploration for posterior segment disease, and that external eye photographs may hold useful data for stratifying disease vs non-disease states. There are several possible explanations for the predictive ability of these models. First, DL models may detect subtle features of the fundus visible within corneal reflections. Second, external structural changes associated with systemic conditions (e.g., diabetic cornea changes, age-related periocular changes, diabetic cataracts) may increase pre-test probability for posterior segment dis-eases. Saliency maps for both DR and AMD frequently highlighted the pupil and the red reflex, as well as the iris-scleral junction, and less frequently the adnexal features. We therefore investigated whether the signal depends on a pharmacologically dilated pupil. In the 857 test images with an undilated pupil, discriminatory power persisted for both vision-threatening DR and advanced AMD, although with lower point estimates and wide confidence intervals (Supplementary Table S17). Previous studies of external eye photographs have found that predictions of DR and systemic biomarkers generalised from undilated to dilated pupils and relied predominantly on the central region of the image (sclera, iris and pupil) rather than the eyelids.[3] These observations argue against the claim that the signal consists exclusively of a low-resolution image of the fundus through a dilated pupil. However, we cannot rule out the possibility that the red reflex contributes part of the signal. To resolve this issue, an explicit experiment involving pupil masking and subsequent retraining is needed.

Consistent with prior work showing that external eye photographs encode systemic biomarkers,[2, 3] all DR models, and the leading AMD and glaucoma models, captured signal significantly beyond a demographics-only baseline. Because this baseline already conditioned on age, race, and ethnicity, the image-specific increment cannot be attributed to these demographic factors alone. This increase was greatest for DR, for which a plausible bio-logical mechanism has been proposed. External eye photographs have been shown to encode information about glycated haemoglobin (HbA1c),[2, 3] and elevated HbA1c is itself an independent risk factor for vision-threatening DR (odds ratio ≈1·33 per 1% increase),[7] so the strong DR signal beyond demographic variables may arise from glycaemia-related oculomic features rather than from demographic information. A comparable, though more tentative, rationale may apply to AMD. Alterations in systemic lipid metabolism have been reported in AMD and linked to disease stage,[8, 10, 9, 32] and external eye photographs can encode circulating markers including blood lipids,[3] so the modest but significant AMD signal beyond demographics may likewise reflect genuine oculomic correlates. We nonetheless present these mechanisms as plausible hypotheses rather than established explanations, and acknowledge that periocular changes correlated with age or other systemic factors may also contribute.

However, it is important to acknowledge that these saliency heat maps do not necessarily explain biological mechanisms for disease classification, but rather give external insight into important pixel-level data that informed its decision. Therefore, a key limitation of our study is that there remains no obviously biological explanations for which external eye photographs may classify posterior segment disease besides the pixel-level information shown and the above hypotheses. Future work on pupil masking and the red re-flex, certain adnexal or extraocular features, or possibly segmentation of the external vascular system may facilitate better understanding of how these models work in predicting posterior segment disease. Ultimately, these findings support the growing body of oculomics literature demonstrating the eye as a window to systemic health.[33]

Of note, it is important to acknowledge that external eye photograph-based models did not outperform fundus photograph-based models; as such, the goal of our study was not to demonstrate that our methodology can or should serve as a screening tool for eye disease, but rather to demonstrate the proof-of-concept utility of using external eye photographs for posterior segment disease classification. Prior DL models for vision-threatening DR from fundus photographs achieved AUCs of 96-97%,[34, 35] and glaucoma models achieved AUCs of 82-91%.[36, 37] Although the image models significantly exceeded a demographics-only baseline for glaucoma (ViT +18·9%, 95% CI 16·3-21·2), absolute discrimination remained modest (AUC 67-74%). This ceiling most plausibly reflects a fundamental constraint of external-eye imaging rather than a shortfall of model capacity or training data: glaucoma is defined by damage to the optic nerve head and retinal nerve fibre layer (posterior structures not visible in external photographs), and its reference standard depends on functional visual-field testing, which need not have any external ocular correlate. This contrasts with diabetic retinopathy and advanced AMD, whose systemic and microvascular determinants (glycaemic and vascular status in DR; ageing- and lipid-related changes in AMD) may leave detectable signatures in the external eye and periocular tissues. Glau-coma may simply lack a comparably strong, externally expressed biomarker.

Our second key finding is that the VLM performed at least as well as the strongest conventional architectures, without statistically significant superiority. LLaVA achieved the highest AUC for DR (85·4% vs 82·6% for ViT), whereas for advanced AMD a convolutional network (EfficientNet, 86·3%) was numerically highest and LLaVA (84·8%) and ViT (84·7%) were comparable, with overlapping 95% confidence intervals across the leading models. This pattern is consistent with prior work reporting comparable performance of VLMs to transformers on healthcare tasks,[38] and high performance of fine-tuned LLaVA on chest X-ray classification.[39] A potential explanation may lie in the optimisation of vision-based tasks using LLaVA’s vision-language connector,[26] which facilitates efficient knowledge transfer from diverse training sources. Our models performed comparably to those published by Babenko et al (AUC 0·79-0·86), despite their larger training cohort.[3] Future work is needed to explore applications of VLMs in other imaging diagnostic applications, as well as novel use cases including image report generation and differential disease generation, especially when used in a multimodal manner (text and imaging).

Our third key finding is that fairness training generally improved overall performance and reduced demographic disparities for DRin single training runs; these comparisons are subject to the caveats on run-to-run variability and minority-group case counts noted in the Limitations. Using ViT as the baseline, most fairness optimisation methods demonstrated comparable or improved ES-AUC compared to baseline training, particularly for race-based equity. These findings were less pronounced for AMD, likely due to significant demographic imbalance in our dataset (87·8% of patients with a positive finding for advanced AMD were White, while in the entire cohort there were only 4 Black patients and 9 Asian patients with a positive AMD finding). The divergent behaviour of race-versus ethnicity-based metrics across DR and AMD (Supplementary Tables S1-S2) likely reflects differing subgroup case mixes and sample sizes: ethnicity was dominated by a large non-Hispanic majority with a small “other” stratum, whereas racial disparities were larger and more systematically reduced by fairness training for DR than for AMD. These results highlight the interplay between data imbalance and algorithmic fairness and the need to account for both dataset and methodological com-ponents of model fine-tuning. Because compilation of representative datasets can be resource-intensive, fairness training methods may offer a complementary approach to promote equitable screening and decrease AI bias against underrepresented demographic groups. We additionally demonstrated simi-lar improvements with FairLoRA in VLMs for DR. To our knowledge, this is a novel finding within AI-based imaging diagnostics and future work should explore how fairness training could potentially improve VLM performance for disease diagnosis within other ophthalmic diseases and other medical specialties as well. Although our experiments were conducted on external eye photographs which are not and should not be used for disease screening with currently existing models, application of these methods for existing AI model applications for posterior segment disease classification may improve model performance and decrease demographic bias, therefore potentially improving model generalization.

Our study has several limitations. First, this was a retrospective study conducted at a single academic center with a predominantly White patient population, which may limit generalisability. Furthermore, 98·7% of the im-ages were acquired on Topcon platforms, so performance on other devices remains unestablished (Supplementary Tables S14-S16). External validation using independent datasets with more diverse populations and acquisition devices is therefore required prior to clinical deployment. Second, disease labels were derived from ICD codes and clinical documentation, which may introduce label noise and misclassification. The comparison groups were de-fined according to the target outcome within the clinical population and were not matched for concomitant ocular or systemic diseases. Therefore, outcome-associated features that are not specific to the target disease, such as cataract or previous ocular surgery, may contribute to the learned signal. Third, we performed binary classification, whereas ophthalmic diseases exist along a clinical spectrum that may differentially affect demographic groups. Fourth, subgroup analyses—particularly for AMD—are limited by small sample sizes in underrepresented racial groups, necessitating cautious interpretation of subgroup-specific performance. Our cohort was 80·1% White, with small minority strata (Black 4·1%, Asian 3·4%, Hispanic 5·6%). Subgroup AUCs in these groups carried very wide confidence intervals, so this imbalance imposes a ceiling on what algorithmic fairness methods can achieve, and apparent fairness gains must be interpreted cautiously and cannot substitute for representative data collection. The overall cohort included only nine Asian patients and four Black patients with advanced AMD. There-fore, the corresponding subgroup AUCs and race-based ES-AUC for AMD should be interpreted with considerable caution, and no claim is made regarding improvements in racial equity for AMD. The observed improvements in race-based ES-AUC for VTDR should likewise be regarded as exploratory findings that require independent replication in larger and more diverse cohorts. Each model configuration was trained once; we did not repeat training across random seeds, so run-to-run variability is not captured by the boot-strap confidence intervals, which quantify sampling variability of the test set only. differences of a few AUC points between methods, such as those observed for sex and ethnicity, may lie within seed-to-seed variation, whereas the race ES-AUC changes for DR (13-19 percentage points for adversarial debiasing, FSCL and FIS) are larger than is typically attributable to seed noise but still derive from small minority-group case counts. Fifth, our study used external eye photographs captured on fundus-camera devices during routine clinical care, and did not assess performance of external photographs acquired by other modalities such as smartphones. While the goal of our proof-of-concept study was to demonstrate feasibility of predicting posterior segment disease, we acknowledge that the performance of our models is in-sufficient for clinical deployment and more work is needed to improve model performance and potentially evaluate on more easily acquired external eye photographs, such as smartphone photographs if the goal is to utilize external photography-based screening for ophthalmic disease.

As a real-world retrospective cohort, the racial and ethnic distribution of our dataset is inherently imbalanced and reflects local patient demographics, referral patterns, and structural access to care. Unlike curated benchmark datasets, clinical datasets cannot be artificially balanced without altering disease prevalence and compromising external validity. Accordingly, it is important to acknowledge that our objective was not to enforce demographic balance, but rather to evaluate model performance and mitigate demographic disparities under realistic clinical data distributions.

We demonstrate that state-of-the-art DL models and VLMs can identify vision-threatening DR and advanced AMD from external eye photographs, signal that significantly exceeds a demographics-only baseline, thereby con-tributing to the growing field of oculomics; the fine-tuned VLM performed comparably to, but not better than, the strongest conventional architectures. We further show that fairness training can improve both overall and demographic-specific performance for DR. These external eye models are a proof-of-concept and are not intended to replace standard-of-care fundus photography or to function as a screening tool in their current form; rather, they establish the feasibility of oculomic analysis of external eye photographs and provide fairness methods that are transferable to other ophthalmic and medical AI applications. Getting closer to clinical readiness would require external and prospective validation in diverse, multi-site populations and acquisition on widely available devices such as smartphones, and patient stakeholders to advance equitable and cost-effective access to care.

## Methods

### Study Design and Ethics

The study was conducted in accordance with the Declaration of Helsinki and approved by the Mass General Brigham Institutional Review Board (approval number: 2024P003332), the institutional review board of record for the Massachusetts Eye and Ear Infirmary, the single participating centre. Because this was a retrospective analysis of existing clinical images and electronic health record data, the Mass General Brigham Institutional Review Board waived the requirement for informed consent. We followed the TRI-POD+AI reporting guidelines for studies developing AI prediction models. There was no patient or public involvement in this study.

### Dataset and Demographics

External eye photographs were obtained from patients who underwent clinical examination at the Massachusetts Eye and Ear Infirmary (MEEI) up until December 2024. The photographs were initially stored with fundus images in a mixed dataset comprising approximately one million images. All images were acquired using a variety of ophthalmic imaging platforms, including Clarus, Visucam, and Visucam NM/FA (Carl Zeiss Meditec); Maestro and Topcon color retinal imaging cameras (Topcon); Silverstone (Optos); and RetCam (Natus Medical). The decision to obtain photographs from a specific machine was determined based on clinician preference and if imaging was felt indicated for documentation purposes or diagnostic assistance. Ophthalmic photographers captured photographs as part of routine clinical care. There was no specific protocol noted for image acquisition, and image resolution, formatting, and metadata varied by the machine used (median resolution 2672×3000 pixels; median field of view 50^◦^; stored in JPEG format; 95·7% of images acquired after pharmacological dilation). Device groups were derived from the device labels contained in the image file names and represent broad imaging platforms rather than confirmed individual camera models. Images labelled Topcon accounted for 98·7% (40,174 of 40,687) of images, with the remainder consisting of images labelled Zeiss (130), RetCam (17) or Optos (2) and unlabelled images (364) (Supplementary Table S14). Patients of all age groups were included; age was defined as the patient’s age at the time the first image for the study was acquired. We employed a three-step procedure to identify external eye photographs.

First, 3,000 randomly sampled images were manually reviewed and categorized into three classes: (1) external eye photographs, (2) fundus images, and (3) unusable images. Unusable images were defined as overexposed im-ages resulting in a completely white image, test images of the device, and images of the room without a human subject. Second, these annotated im-ages were used to train a previously developed deep learning classifier based on the EfficientNet architecture, which was subsequently applied to identify candidate external eye photographs. Third, four trained medical researchers independently reviewed all candidate images and classified them as dilated external photographs, undilated external photographs, non-external photographs, unsure, or unusable. Images classified as “unsure” by a reviewer were subsequently assessed by a senior grader for final adjudication. Unusable images were external eye photographs (not fundus images) that were unsuitable for analysis; reasons for unusability included fully closed eyelids, a non-visible iris, previous keratoprosthesis (KPro) surgery, the presence of corneal fluorescein dye, and acquisition under a non-standard illumination filter. This process resulted in a final dataset of 40,687 external eye photographs from 9,878 unique patients.

Demographic variables including sex, race, ethnicity, and primary language were extracted from the electronic health record. Self-reported sex was defined as male and female. Race and ethnicity were self-reported by patients at registration. Race categories included Asian, Black or African American, White, and Other (including multiple races and unknown). Ethnicity was categorised as Hispanic, non-Hispanic, or unknown.

### Disease Definitions

Disease labels were obtained from clinical diagnoses and the electronic health record as follows:

AMD diagnosis was based on International Classification of Diseases (ICD-10) codes. Subjects were categorised into four classes: normal, early AMD, intermediate AMD, and advanced AMD (including both geographic atrophy and neovascular AMD). For this study, we performed binary classification of advanced AMD versus all other categories. ICD-10 codes have been assigned by treating ophthalmologist and DR severity followed the International Clinical Diabetic Retinopathy Severity Scale.

DR diagnosis was based on ICD-10 codes. Subjects were classified as having non-vision-threatening DR (no DR, mild non-proliferative DR, or moder-ate non-proliferative DR) or vision-threatening DR (severe non-proliferative DR or proliferative DR). For both DR and AMD, the comparison group therefore comprised all other patients in the cohort, including those with less severe stages of the same disease and those with other ocular diagnoses, rather than healthy controls. Comparison groups were not matched for concomitant ocular disease, because the aim was to detect advanced disease within a real-world clinical population rather than to isolate disease-specific features under a matched case-control design.

Due to variability in clinician input ICD codes across patients, glaucoma diagnosis was defined based on visual field metrics rather than ICD codes. Subjects were categorised as normal (visual field mean deviation ≥ −1 dB and otherwise relatively normal pattern standard deviation) or glaucoma (visual field mean deviation *<* −3 dB with abnormal pattern standard deviation).

### Data Partitioning

Data were split at the patient level to prevent data leakage, with no images from the same patient appearing in both training and test sets. The split was stratified by disease status to maintain representative distributions. The dataset was divided into training (n=20,307 images from 4,939 patients) and test (n=20,380 images from 4,939 patients) sets at approximately 1:1 ratio. No separate validation set was used for hyperparameter tuning.

### Model Architectures

We evaluated three widely adopted computer vision architectures: WRes-Net (specifically Wide-ResNet-38), EfficientNet (specifically EfficientNet-B5), and ViT (DeiT-Small, distilled). We additionally fine-tuned a vision-language model, LLaVA-7B, using Low-Rank Adaptation (LoRA) to preserve pre-trained representations while enabling domain-specific adaptation (Table 2). For the LLaVA model, a structured multimodal prompt was used (Box 1), consisting of a system instruction and a user query paired with the input image. The system instruction constrained the model to provide a binary diagnostic output without explanation, while the user query requested disease classification based on the image. The model was fine-tuned with LoRA under a standard autoregressive (language-modelling) cross-entropy loss on the answer token. For evaluation, the discrete generated token was not used directly; instead, at the answer position we took the logits of the “positive” and “negative” tokens and computed *p* = *σ*(*ℓ*_positive_ − *ℓ*_negative_) ∈ [0, 1], the model’s probability of “positive” relative to “negative”, where *σ* denotes the sigmoid function. This continuous score was used to compute AUC and ESAUC, consistent with the other models. The full prompt template is shown in Box 1.

**Table 2:** Model architectures and training hyperparameters. LoRA = Low-Rank Adaptation; *r* = LoRA rank; *α* = LoRA scaling factor.

| Model | Batch Size | Epochs | Input Size | Learning Rate | Variant | Additional |
|---|---|---|---|---|---|---|
| WResNet | 16 | 20 | 448 | 5e-3 | WResNet-38 | – |
| EfficientNet | 32 | 20 | 456 | 1e-3 | EfficientNet-B5 | – |
| ViT | 64 | 60 | 448 | 5e-4 | DeiT-S (Small) | Distilled |
| LLaVA | 32 | 15 | 336 | 2e-4 | LLaVA-7B | LoRA *r* = 8, *α* = 16 |

### Image Preprocessing and Data Augmentation

All deep learning models were initialised with ImageNet-pretrained weights, and inputs were normalised using standard ImageNet statistics. During training, images were augmented with *RandomResizedCrop*, random horizontal flipping (probability 0·5), and colour jittering (brightness, contrast, and saturation 0·4; hue 0·1). For ViT, we additionally applied the AutoAug-ment policy to improve robustness. Input resolution was 448×448 for ViT and WResNet, 456×456 for EfficientNet, and 336×336 for LLaVA. The ex-act ImageNet normalisation parameters were a mean of [0·485, 0·456, 0·406] and a standard deviation of [0·229, 0·224, 0·225] across the red, green, and blue channels. All experiments were performed in Python 3.9.21 and Py-Torch 2.5.0, with computation accelerated on an NVIDIA RTX A6000 GPU (CUDA 12.4).

#### Box 1 Prompt template used for LLaVA fine-tuning and inference

The system instruction and user query shown below were paired with each external eye photograph. The placeholder >DISEASE_NANE> was replaced by the target condition (vision-threatening diabetic retinopathy, advanced age-related macular degeneration, or glaucoma).

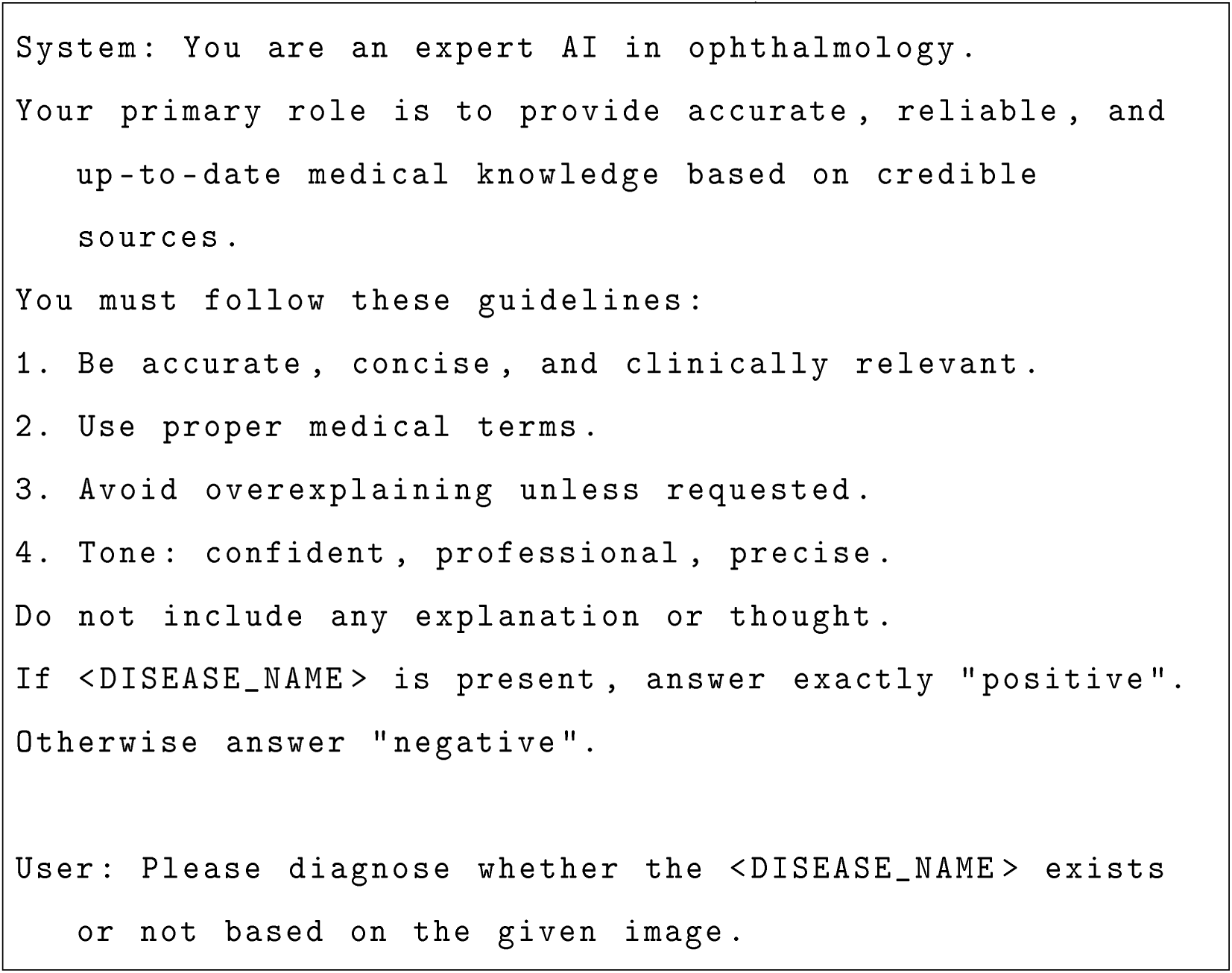

### Training and Hyperparameter Tuning

ViT was trained using the AdamW optimiser and the convolutional models (WResNet, EfficientNet) using stochastic gradient descent, all with bi-nary cross-entropy loss. ViT was trained for 60 epochs, the convolutional models for 20 epochs, and LLaVA was fine-tuned for 15 epochs. Learning rates, batch sizes and training epochs were predetermined based on previous experiments with these architectures, without using the held-out test set for hyperparameter optimisation. The final learning rates and batch sizes were: WResNet (5×10^−3^, 16), EfficientNet (1×10^−3^, 32), ViT (5×10^−4^, 64) and LLaVA (2×10^−4^, 32) (Table 2). All fairness-trained variants inherited the optimisation settings of their corresponding baseline architectures.

Fairness-specific hyperparameters were adapted from previously published studies[21, 31] and fixed before evaluation on the held-out test set. The test set was used exclusively for final performance evaluation.

### Pairness Optimisation Methods

We evaluated five fairness-enhancing methods:

1. Distributionally robust optimisation (DRO) improves fairness by modeling data uncertainty without prior knowledge of data distribution. Prior work has shown that in overparameterized models, Grouped DRO alone offers little benefit over Empirical Risk Minimization (ERM), and that improvements in worst-group performance arise when the optimization is conducted with strong regularization techniques. [27]
2. Adversarial debiasing (Adv) is an adversarial training-based fairness approach that removes sensitive attribute information from latent representations. A task classifier and an adversarial sensitive-attribute classifier are jointly trained, where gradients from the sensitive classifier are reversed to discourage sensitive information in the learned representation.[28]
3. Fair supervised contrastive loss (FSCL) addresses the tendency of supervised contrastive learning to encode sensitive-attribute information. FSCL mitigates this by restricting the negative set to samples that share the same sensitive attribute but belong to a different target class, so that the representation is not encouraged to separate classes in a way that relies on the sensitive attribute.[29]
4. Fair identity scaling (FIS) is a fairness approach tailored to medical AI, which views fairness as both a group-level and identity-level problem. It proposes an identity-level scaling mechanism that downweights patients the model already performs well on, while emphasizing patients with poorer generalization, who often belong to minority groups. Further-more, it applies normalization across demographic groups in addition to identity-level scaling to prevent imbalances between groups.[30]
5. FairLoRA addresses group fairness in federated learning by extending Low-Rank Adaptation (LoRA) with a fairness-aware variant. By interpreting low-rank updates from an SVD perspective, the method shares disease-related common visual patterns across all groups while learning group-specific scaling factors that capture differing feature importance for each demographic group.[31]

DRO, Adv, FSCL, and FIS were applied to ViT; FairLoRA was applied to LLaVA. The mathematical formulations and training procedures for each method are detailed below.

The four ViT-based fairness methods (DRO, Adv, FSCL, and FIS) operated on the ViT backbone (DeiT-S, embedding dimension 384). Each was trained for one protected attribute *s* ∈ *S* at a time (sex, race, or ethnicity), with group cardinality |*S*|. For adversarial debiasing and FSCL, the relevant representation is the final-block patch-token feature map *F* ∈ ℝ^384×*H*′×*W*′^ (before the classification head); DRO and FIS act only on the loss.

For DRO, with per-sample binary cross-entropy losses, let *L_g_* be the mean loss of group *g*. Group weights *q_g_* (initialised to 1*/*|*S*|) are updated multiplicatively from the normalised group losses 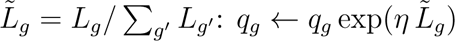, renormalised to sum to one, with step size *η* = 0.01; the training loss is the worst-group-weighted sum ∑*_g_ q_g_L_g_*.

For adversarial debiasing, an auxiliary group classifier *g_ψ_* (a 3×3 convolution 384 →|*S*| followed by global average pooling) is attached to *F* through a gradient-reversal layer *R_λ_* that is the identity in the forward pass and negates the gradient in the backward pass, *∂R_λ_*(*F*)*/∂F* = −*λI*, with *λ* = 1.0. The objective is *L* = *L*_cls_ + *α L*_CE_(*g_ψ_*(*R_λ_*(*F*))*, s*), *α* = 0.01, so the auxiliary head learns to predict *s* while the reversed gradient drives the backbone to make *F* uninformative of *s*.

For FSCL, a projection head maps *F* to an *ℓ*_2_-normalised embedding *z* ∈ ℝ^64^. For each anchor *i*, the positives *P* (*i*) are its two augmented views and the contrastive denominator is restricted to samples sharing the same sensitive attribute:

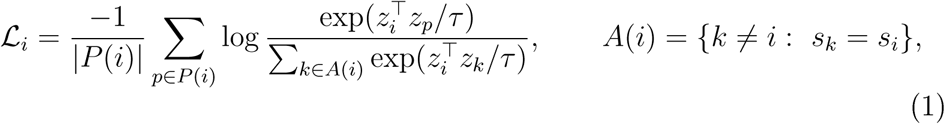

with temperature *τ* = 0.1. Training is two-stage: contrastive representation learning, followed by classifier fine-tuning with binary cross-entropy.

FIS reweights the per-sample classification loss so that training focuses on under-served samples. Two weights are combined: an individual weight that emphasises samples on which the model performs poorly (high loss), and a group weight that emphasises demographic groups whose loss distribution deviates most from the overall distribution. Concretely, for per-sample losses *ℓ*, *w*^ind^ = *ρ* softmax(*ℓ/T*) upweights high-loss samples, and *w*^grp^ = *ρ* softmax(*d_s_/T*) upweights groups with large *d_g_*, the Sinkhorn distance (a distributional distance; entropic optimal transport, *p*=2, blur 0.1) between group *g*’s loss distribution and the pooled distribution. The final weight *w* = (1 − *β*) *w*^ind^ + *β w*^grp^ yields the FIS loss, with *T* = 1.0, *ρ* = 2.0, and *β* = 0.5; the total objective is *L*_cls_ + 10 *L*_FIS_.

To analyse the effect of fairness-oriented fine-tuning on VLMs, we adopted FairLoRA, which integrates fairness-aware training with LoRA. LoRA enables fine-tuning with a reduced number of trainable parameters by adding trainable low-rank parameters to the pre-trained weights. Specifically, for a pre-trained weight matrix *W*_0_ ∈ ℝ*^m^*^×^*^n^*, the fine-tuned weight is written as

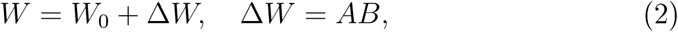

where *A* ∈ ℝ*^m^*^×^*^r^* and *B* ∈ ℝ*^r^*^×^*^n^*, with small rank *r*. The resulting low-rank update admits an equivalent formulation via singular value decomposition (SVD),

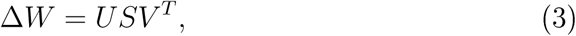

where *U* ∈ ℝ*^m^*^×^*^r^* and *V* ∈ ℝ*^n^*^×^*^r^* are the left and right singular vector matrices and *S* ∈ ℝ*^r^*^×^*^r^* is a diagonal singular value matrix; the singular vectors capture global structural directions across the embedding space, while the singular values control the relative importance of each direction. FairLoRA extends this framework to incorporate group fairness by disentangling shared and group-specific components of the low-rank update. For a demographic group *g*, FairLoRA defines the LoRA weights as

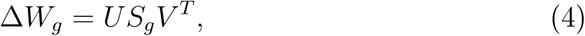

where *U* and *V* are shared across all demographic groups (capturing inter-group relationships) and *S_g_* is a group-specific singular value matrix (pre-serving intra-group distinctiveness). In this study, we applied FairLoRA to LLaVA-7B with LoRA rank *r* = 8 and scaling factor *α* = 16 to evaluate the impact of fairness-aware fine-tuning on VLM performance.

### Outcome Measures

The primary outcome was the AUC for disease classification for each model and disease. Secondary outcomes included sensitivity at fixed specificity thresholds (80%, 85%, 90%, 95%) and equity-scaled AUC (ES-AUC) for each demographic group across each model. ES-AUC was calculated as:

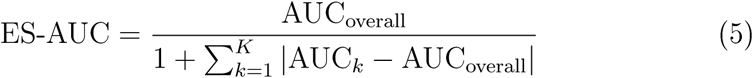

where AUC_overall_ (scale 0-1) denotes the overall test-set AUC, and AUC*_k_* denotes the subgroup AUC for demographic group *k*. This metric preserves overall discriminative performance while penalizing larger deviations between subgroup AUCs and the overall AUC.

Three units of analysis were used and are stated explicitly here: the dataset was partitioned by patient; AUC, sensitivity and ES-AUC were computed over test-set images; and prevalence is reported per patient. Uncertainty was quantified by patient-clustered bootstrap resampling of the test set: in each of 2,000 iterations, patient identifiers were sampled with replacement and all eligible images of each sampled patient were retained together (and duplicated when a patient was drawn more than once), so that within-patient correlation between images is reflected in the interval width. We report 95% confidence intervals as the 2·5th and 97·5th percentiles of the bootstrap distribution for AUC, sensitivity at fixed specificity, subgroup AUC and ES-AUC; maximum subgroup disparity is reported as a point estimate. Because ES-AUC is a function of absolute deviations between subgroup and overall AUCs, its bootstrap distribution is shifted towards lower values than the full-sample estimate (resampling inflates the subgroup deviations, particularly in small strata), so ES-AUC intervals are asymmetric and the point estimate typically lies in the upper part of the interval. The paired com-parisons against the demographics-only baseline and between each fairness method and its baseline (Supplementary Tables S8-S10) were computed with 100 patient-clustered resamples and paired Student’s t-tests, with statistical significance defined as p *<* 0·05; these paired analyses were not recomputed with 2,000 resamples. Bootstrap intervals quantify sampling variability of the test set only and do not capture run-to-run variability of model training, which was performed once per configuration.

## Data Availability

The data supporting the findings of this study are not publicly available due to patient privacy constraints. The dataset includes external eye photographs containing identifiable facial features that preclude open sharing. All data are stored in a controlled-access repository at Massachusetts Eye and Ear. This study was conducted under a protocol approved by the Mass General Brigham Institutional Review Board (approval number: 2024P003332).

## Code Availability

The code for model training and evaluation is publicly available in the Harvard Ophthalmology AI Lab GitHub repository at https://github.com/Harvard-AI-and-Robotics-Lab/FairExternalEye.

## Acknowledgements

We thank the patients of the Massachusetts Eye and Ear Infirmary whose data enabled this research.

## Author Contributions

Author initials: VK (Vasil Kostin), SY (Sung-Hoon Yoon), JSC (Jimmy S. Chen), SB (Seungjoo Baek), JS (Jesse Seid), NO (Nathan Ouyang), EdL (Edward Li), ErL (Eric Li), MS (Min Shi), and MW (Mengyu Wang). VK, SY, and JSC contributed equally to this work and should be considered co-first authors.

**Conceptualization**: VK, MS, MW. **Data curation**: VK, MS, SY, JS, NO, EdL, ErL. **Formal analysis**: VK, SY, MS. **Funding acquisition:** MW, MS. **Investigation**: VK, SY, JSC, MS, JS, NO, EdL, ErL. **Methodology**: VK, SY, MW, MS. **Project administration**: VK, SY, MW. **Software**: SY, VK, MS. **Supervision**: MW. **Validation**: SY, VK, JSC, MW. **Visualization**: VK, SY, SB. **Writing — original draft:** VK, JSC, SY. **Writing — review & editing**: All authors. All authors read and approved the final version of the manuscript.

All authors had full access to all the data in the study and had final responsibility for the decision to submit for publication. VK, SY, MS, and SB directly accessed and verified the underlying data reported in the manuscript.

