## Supplementary Materials for "Applications of Fairness Training in Artificial Intelligence to Ophthalmic Disease Diagnosis using External Eye Photographs"

[*] These authors contributed equally to this manuscript

#### Contents

**1 Supplementary Tables** **2**

### 1 Supplementary Tables

#### Fairness-training results, baseline discrimination, and equity (Tables S1–S4)

Supplementary Tables S1–S4 report the per-disease fairness-training results (Table S1, diabetic retinopathy; Table S2, advanced AMD), baseline model discrimination (Table S3), and baseline equity-scaled AUC (Table S4). AUCs and ES-AUC values are shown with 95% bootstrap confidence intervals (2,000 patient-clustered resamples of the test set); the maximum between-subgroup disparity is a point estimate.

**Table S1: Fairness training results for vision-threatening diabetic retinopathy.** AUCs and ES-AUC with 95% bootstrap CIs (2,000 resamples); ES-AUC = equity-scaled AUC; Max Disp. = maximum between-subgroup AUC difference (point estimate). LLaVA + FairLoRA values are point estimates without CIs. Best value per column in **bold**.

**Sex**

| Method | Overall | ES-AUC | Max Disp. | Female | Male |
|---|---|---|---|---|---|
| ViT (baseline) | 82.6 (79.9, 85.1) | 81.4 (76.5, 84.1) | 1.5% | 81.8 (77.9, 85.4) | 83.3 (79.7, 86.9) |
| ViT + DRO | 78.4 (75.2, 81.2) | 75.9 (70.4, 79.8) | 3.2% | 79.8 (75.5, 83.8) | 76.6 (72.0, 80.9) |
| ViT + Adv | 84.8 (82.2, 87.2) | 84.7 (79.1, 86.3) | **0.1%** | 84.7 (80.9, 88.2) | 84.8 (80.8, 88.1) |
| ViT + FSCL | 83.0 (80.3, 85.5) | 82.5 (76.8, 84.6) | 0.7% | 83.3 (79.7, 86.9) | 82.6 (78.5, 86.3) |
| ViT + FIS | **85.7 (83.1, 88.1)** | **85.4 (79.5, 87.2)** | 0.4% | **85.4 (81.1, 89.2)** | **85.8 (82.3, 89.0)** |
| LLaVA + FairLoRA | 83.2 | 83.0 | 0.3% | 83.1 | 83.4 |

**Race**

| Method | Overall | ES-AUC | Max Disp. | Asian | Black | White | Other |
|---|---|---|---|---|---|---|---|
| ViT (baseline) | 82.6 (79.9, 85.1) | 56.9 (49.7, 64.8) | 25.9% | 56.7 (39.0, 74.2) | 70.5 (63.3, 77.0) | 75.4 (70.8, 80.0) | 82.6 (78.6, 86.5) |
| ViT + DRO | 81.0 (78.1, 83.8) | 60.7 (54.5, 66.4) | 13.5% | 66.6 (54.6, 77.4) | 68.9 (60.3, 76.6) | 75.1 (69.6, 80.0) | 80.1 (74.6, 85.1) |
| ViT + Adv | 83.4 (80.3, 86.3) | **75.6 (64.0, 77.0)** | **5.6%** | 83.1 (69.0, 95.9) | **79.1 (72.3, 85.0)** | 78.1 (73.7, 82.6) | **83.7 (77.7, 89.3)** |
| ViT + FSCL | 84.1 (81.3, 86.5) | 73.3 (65.0, 77.0) | 10.4% | 85.3 (75.5, 93.9) | 74.9 (66.4, 83.0) | 80.8 (76.5, 84.9) | 83.0 (77.7, 87.6) |
| ViT + FIS | 84.2 (81.3, 86.7) | 69.9 (63.0, 74.6) | 13.1% | **87.3 (78.2, 95.4)** | 74.2 (66.8, 80.4) | 79.8 (75.8, 83.6) | 81.3 (74.8, 86.7) |
| LLaVA + FairLoRA | **86.3** | 73.0 | 10.4% | 85.6 | 75.2 | **83.1** | 83.1 |

**Ethnicity**

| Method | Overall | ES-AUC | Max Disp. | Non-Hisp | Hispanic | Other |
|---|---|---|---|---|---|---|
| ViT (baseline) | 82.6 (79.9, 85.1) | 64.3 (54.4, 77.1) | 28.3% | 82.5 (79.8, 85.1) | 85.6 (80.3, 90.0) | 57.3 (36.1, 80.2) |
| ViT + DRO | 83.2 (80.4, 85.7) | 64.4 (54.4, 77.4) | 29.1% | 83.4 (80.6, 86.1) | 85.7 (79.5, 90.5) | 56.6 (35.9, 79.2) |
| ViT + Adv | 84.0 (81.1, 86.7) | 64.1 (53.1, 78.5) | 30.5% | 84.6 (81.7, 87.2) | **86.3 (77.6, 92.6)** | 55.8 (32.4, 80.0) |
| ViT + FSCL | 83.3 (80.6, 85.8) | 66.2 (54.7, 78.5) | 23.8% | 84.1 (81.5, 86.6) | 81.3 (69.9, 90.3) | 60.3 (37.4, 83.1) |
| ViT + FIS | **84.3 (81.5, 86.9)** | 67.2 (55.0, 79.3) | 25.3% | **85.2 (82.3, 87.7)** | 84.0 (74.0, 91.3) | 59.9 (37.9, 83.2) |
| LLaVA + FairLoRA | 83.7 | **72.4** | **15.2%** | 84.1 | 85.0 | **69.8** |

**Language**

| Method | Overall | ES-AUC | Max Disp. | English | Spanish | Other |
|---|---|---|---|---|---|---|
| ViT (baseline) | 82.6 (79.9, 85.1) | 70.9 (61.4, 78.4) | 15.7% | 81.8 (78.8, 84.6) | 89.5 (85.8, 92.7) | 73.8 (56.7, 90.2) |
| ViT + DRO | 82.3 (79.6, 84.8) | 70.1 (60.2, 77.2) | 16.4% | 81.3 (78.2, 84.2) | 90.6 (86.8, 93.8) | 74.2 (56.3, 90.5) |
| ViT + Adv | 83.7 (80.7, 86.4) | 71.7 (61.5, 76.4) | 15.0% | 82.0 (78.6, 85.0) | **95.2 (92.8, 97.3)** | 80.2 (62.5, 93.2) |
| ViT + FSCL | 82.5 (79.6, 85.1) | 69.5 (60.5, 75.0) | 17.1% | 80.9 (77.7, 83.9) | 94.5 (92.0, 96.9) | 77.4 (61.2, 91.7) |
| ViT + FIS | 84.3 (81.5, 86.8) | **73.0 (63.9, 77.9)** | 14.2% | 83.1 (80.0, 86.0) | 94.3 (91.7, 96.7) | 80.1 (65.0, 92.2) |
| LLaVA + FairLoRA | **85.5** | 72.7 | **10.3%** | **83.6** | 93.9 | **92.8** |

**Table S2: Fairness training results for advanced age-related macular degeneration.** AUCs and ES-AUC with 95% bootstrap CIs (2,000 resamples); ES-AUC = equity-scaled AUC; Max Disp. = maximum between-subgroup AUC difference (point estimate). LLaVA + FairLoRA values are point estimates without CIs. Best value per column in **bold**.

**Sex**

| Method | Overall | ES-AUC | Max Disp. | Female | Male |
|---|---|---|---|---|---|
| ViT (baseline) | 84.7 (82.8, 86.5) | 81.8 (78.3, 85.1) | 3.5% | 83.3 (80.8, 85.8) | 86.8 (84.3, 89.2) |
| ViT + DRO | 83.7 (81.8, 85.5) | 79.7 (76.3, 83.4) | 5.0% | 81.7 (79.1, 84.4) | 86.7 (84.3, 89.0) |
| ViT + Adv | 84.4 (82.5, 86.3) | 80.6 (77.2, 84.3) | 4.8% | 82.4 (79.9, 85.0) | 87.2 (84.6, 89.6) |
| ViT + FSCL | **85.4 (83.6, 87.2)** | **81.9 (78.4, 85.3)** | 4.3% | **83.6 (81.1, 86.0)** | **87.9 (85.3, 90.5)** |
| ViT + FIS | 74.4 (71.9, 77.1) | 72.7 (68.4, 75.9) | **2.4%** | 73.8 (70.6, 77.2) | 76.2 (72.3, 79.9) |
| LLaVA + FairLoRA | 83.9 | 79.5 | 5.5% | 81.8 | 87.3 |

**Race**

| Method | Overall | ES-AUC | Max Disp. | Asian | Black | White | Other |
|---|---|---|---|---|---|---|---|
| ViT (baseline) | 84.7 (82.8, 86.5) | 60.5 (48.6, 77.7) | 35.6% | 62.2 (29.8, 93.7) | 97.8 (94.9, 100.0) | 85.0 (82.9, 86.9) | **80.5 (75.7, 85.2)** |
| ViT + DRO | 83.0 (80.9, 85.0) | **67.5 (61.0, 79.4)** | **21.6%** | **83.9 (80.5, 87.2)** | 97.4 (94.3, 99.7) | 83.4 (81.3, 85.4) | 75.8 (66.7, 84.3) |
| ViT + Adv | 85.0 (83.1, 86.9) | 61.4 (53.1, 79.8) | 32.0% | 67.3 (47.1, 86.8) | 99.3 (98.6, 99.9) | 85.4 (83.4, 87.4) | 79.1 (73.1, 85.1) |
| ViT + FSCL | **85.2 (83.2, 87.0)** | 55.7 (46.5, 75.3) | 42.7% | 55.9 (28.4, 83.0) | 98.6 (97.2, 99.7) | **86.1 (84.2, 88.0)** | 76.0 (68.8, 82.9) |
| ViT + FIS | 84.4 (82.5, 86.2) | 56.6 (45.6, 80.3) | 44.9% | 54.6 (21.6, 87.2) | **99.5 (99.0, 100.0)** | 84.6 (82.6, 86.4) | 80.4 (73.3, 87.1) |
| LLaVA + FairLoRA | 83.5 | 61.6 | 29.3% | 62.1 | 91.4 | 84.6 | 78.4 |

**Ethnicity**

| Method | Overall | ES-AUC | Max Disp. | Non-Hisp | Hispanic | Other |
|---|---|---|---|---|---|---|
| ViT (baseline) | 84.7 (82.8, 86.5) | **79.7 (70.6, 83.2)** | **5.5%** | 84.4 (82.4, 86.3) | 89.9 (81.5, 96.8) | 85.4 (75.1, 93.2) |
| ViT + DRO | 83.8 (81.8, 85.7) | 78.8 (69.4, 82.8) | 6.2% | 83.6 (81.5, 85.6) | 87.3 (76.5, 95.8) | 81.1 (71.6, 89.6) |
| ViT + Adv | 84.3 (82.2, 86.2) | 77.4 (66.3, 83.0) | 8.8% | 84.2 (82.0, 86.2) | 88.1 (77.5, 96.8) | 79.3 (65.4, 91.9) |
| ViT + FSCL | **85.9 (84.1, 87.6)** | 70.3 (59.8, 78.3) | 21.7% | **86.3 (84.5, 88.1)** | 85.6 (74.7, 95.1) | 64.6 (48.5, 79.5) |
| ViT + FIS | 84.9 (83.1, 86.7) | 77.8 (69.5, 83.1) | 6.7% | 84.5 (82.5, 86.3) | **91.2 (80.0, 98.8)** | **87.4 (78.8, 94.8)** |
| LLaVA + FairLoRA | 81.9 | 67.1 | 21.8% | 81.7 | 91.1 | 69.3 |

**Language**

| Method | Overall | ES-AUC | Max Disp. | English | Spanish | Other |
|---|---|---|---|---|---|---|
| ViT (baseline) | 84.7 (82.8, 86.5) | 74.9 (69.3, 81.3) | 8.4% | 84.1 (82.1, 86.0) | 92.5 (85.0, 98.4) | 89.3 (84.5, 93.7) |
| ViT + DRO | 84.5 (82.6, 86.4) | **77.7 (71.6, 82.9)** | **6.1%** | 84.1 (82.1, 86.1) | 90.2 (83.2, 95.5) | 87.1 (81.2, 92.1) |
| ViT + Adv | 84.5 (82.5, 86.5) | 75.7 (69.7, 82.0) | 8.4% | 84.1 (81.8, 86.1) | 92.5 (85.2, 98.2) | 87.9 (82.2, 92.9) |
| ViT + FSCL | **85.6 (83.8, 87.4)** | 75.1 (70.0, 81.0) | 8.6% | **85.0 (83.0, 86.9)** | 93.6 (88.2, 97.9) | **91.0 (85.9, 95.1)** |
| ViT + FIS | 84.3 (82.4, 86.1) | 73.9 (68.6, 79.0) | 10.5% | 83.7 (81.6, 85.6) | **94.2 (89.9, 97.5)** | 88.0 (82.4, 92.9) |
| LLaVA + FairLoRA | 82.1 | 73.4 | 7.2% | 82.2 | 89.4 | 86.5 |

**Table S3:** Baseline model discrimination: AUC (95% CI; 2,000 bootstrap resamples) and sensitivity (%) at fixed specificity. LLaVA values are point estimates without CIs. Glaucoma was excluded from fairness analysis.

| Disease | Model | AUC | Sens@80 | Sens@85 | Sens@90 | Sens@95 |
|---|---|---|---|---|---|---|
| DR | WResNet | 78.2 (74.9, 81.4) | 64.2 (57.5, 71.0) | 54.9 (47.8, 61.7) | 43.5 (36.4, 50.7) | 26.9 (20.1, 32.6) |
| | ViT | 82.6 (79.9, 85.1) | 66.3 (59.6, 72.8) | 60.1 (52.9, 66.8) | 49.7 (42.7, 56.8) | 34.7 (27.6, 41.3) |
| | EfficientNet | 78.7 (75.3, 81.8) | 61.7 (54.7, 68.5) | 55.4 (48.1, 62.4) | 46.1 (38.9, 53.0) | 28.0 (22.1, 34.7) |
| | LLaVA | 85.4 | 76.7 | 71.5 | 61.7 | 47.2 |
| Advanced AMD | WResNet | 83.0 (80.9, 85.0) | 70.2 (65.4, 75.3) | 60.5 (55.5, 65.8) | 47.4 (41.9, 52.7) | 32.7 (27.7, 38.2) |
| | ViT | 84.7 (82.8, 86.5) | 73.9 (69.3, 78.8) | 64.5 (58.9, 69.5) | 51.4 (46.2, 56.7) | 34.7 (29.9, 40.0) |
| | EfficientNet | 86.3 (84.5, 88.1) | 77.3 (73.0, 81.7) | 67.0 (62.0, 71.9) | 57.7 (52.6, 63.0) | 44.0 (38.4, 49.2) |
| | LLaVA | 84.8 | 73.3 | 65.1 | 55.1 | 36.4 |
| Glaucoma | WResNet | 67.5 (65.5, 69.5) | 41.5 (37.9, 45.1) | 35.1 (31.5, 38.7) | 27.2 (24.0, 30.7) | 16.8 (14.2, 19.7) |
| | ViT | 73.9 (71.9, 75.8) | 52.2 (48.4, 56.2) | 46.1 (42.5, 50.1) | 37.4 (33.8, 41.2) | 26.7 (23.5, 30.3) |
| | EfficientNet | 67.7 (65.7, 69.6) | 41.4 (37.8, 45.2) | 33.2 (29.8, 37.1) | 25.0 (22.1, 28.5) | 15.1 (12.5, 17.9) |

**Table S4:** Baseline equity-scaled AUC (ES-AUC, %) by demographic attribute, with 95% bootstrap CIs (2,000 resamples). LLaVA values are point estimates without CIs.

| Disease | Model | ES-AUC (Sex) | ES-AUC (Race) | ES-AUC (Ethnicity) | ES-AUC (Language) |
|---|---|---|---|---|---|
| DR | WResNet | 75.1 (68.9, 80.2) | 61.0 (54.0, 67.0) | 64.7 (51.3, 75.7) | 67.6 (59.4, 74.7) |
| | ViT | 81.4 (76.5, 84.1) | 56.9 (49.7, 64.8) | 64.3 (54.4, 77.1) | 70.9 (61.4, 78.4) |
| | EfficientNet | 74.7 (68.9, 79.9) | 66.1 (58.4, 70.7) | 73.4 (62.0, 77.2) | 69.4 (58.5, 76.3) |
| | LLaVA | 83.5 | 71.9 | 67.6 | 71.3 |
| Advanced AMD | WResNet | 77.3 (73.7, 81.0) | 51.5 (48.2, 72.5) | 71.9 (62.6, 78.6) | 70.7 (65.3, 75.6) |
| | ViT | 81.8 (78.3, 85.1) | 60.5 (48.6, 77.7) | 79.7 (70.6, 83.2) | 74.9 (69.3, 81.3) |
| | EfficientNet | 80.3 (76.8, 84.0) | 63.1 (59.4, 76.1) | 77.6 (66.0, 83.2) | 78.2 (71.9, 80.7) |
| | LLaVA | 81.1 | 61.2 | 75.8 | 79.5 |
| Glaucoma | WResNet | 65.1 (62.0, 68.1) | 50.2 (44.4, 55.5) | 54.3 (49.1, 59.7) | 55.0 (49.9, 57.7) |
| | ViT | 72.0 (68.8, 74.8) | 54.9 (48.4, 61.1) | 60.9 (55.6, 67.2) | 61.9 (57.2, 66.3) |
| | EfficientNet | 67.4 (64.0, 68.9) | 47.7 (42.9, 51.8) | 56.1 (51.0, 60.4) | 55.1 (50.4, 57.9) |

**Table S5: Baseline model performance for vision-threatening diabetic retinopathy (no fairness training).** AUCs and ES-AUC with 95% bootstrap CIs (2,000 resamples); Max Disp. = max between-subgroup AUC difference (point). LLaVA values are point estimates without CIs. Best per column **bold**.

| **Sex** | | | | | |
|---|---|---|---|---|---|
| Model | Overall | ES-AUC | Max Disp. | Female | Male |
| WResNet | 78.2 (74.9, 81.4) | 75.1 (68.9, 80.2) | 4.2% | 76.0 (70.6, 81.3) | 80.2 (76.1, 84.0) |
| ViT | 82.6 (79.9, 85.1) | 81.4 (76.5, 84.1) | **1.5%** | 81.8 (77.9, 85.4) | 83.3 (79.7, 86.9) |
| EfficientNet | 78.7 (75.3, 81.8) | 74.7 (68.9, 79.9) | 5.3% | 75.8 (70.9, 80.7) | 81.1 (76.9, 84.8) |
| LLaVA | **85.4** | **83.5** | 2.3% | **86.5** | **84.2** |

| **Race** | | | | | | | |
|---|---|---|---|---|---|---|---|
| Model | Overall | ES-AUC | Max Disp. | Asian | Black | White | Other |
| WResNet | 78.2 (74.9, 81.4) | 61.0 (54.0, 67.0) | 17.9% | 71.5 (60.8, 83.6) | 62.5 (53.1, 71.8) | 74.5 (69.5, 79.2) | 80.4 (74.4, 85.6) |
| ViT | 82.6 (79.9, 85.1) | 56.9 (49.7, 64.8) | 25.9% | 56.7 (39.0, 74.2) | 70.5 (63.3, 77.0) | 75.4 (70.8, 80.0) | 82.6 (78.6, 86.5) |
| EfficientNet | 78.7 (75.3, 81.8) | 66.1 (58.4, 70.7) | **5.4%** | 74.0 (60.8, 87.4) | 70.6 (61.5, 79.7) | 76.0 (70.9, 80.6) | 75.1 (68.8, 81.0) |
| LLaVA | **85.4** | **71.9** | 13.4% | **93.3** | **79.9** | **80.1** | **85.4** |

| **Ethnicity** | | | | | | |
|---|---|---|---|---|---|---|
| Model | Overall | ES-AUC | Max Disp. | Non-Hisp | Hispanic | Other |
| WResNet | 78.2 (74.9, 81.4) | 64.7 (51.3, 75.7) | 15.8% | 79.0 (75.7, 82.2) | 73.1 (61.0, 83.1) | 63.2 (35.2, 90.4) |
| ViT | 82.6 (79.9, 85.1) | 64.3 (54.4, 77.1) | 28.3% | 82.5 (79.8, 85.1) | 85.6 (80.3, 90.0) | 57.3 (36.1, 80.2) |
| EfficientNet | 78.7 (75.3, 81.8) | **73.4 (62.0, 77.2)** | **3.4%** | 78.4 (74.7, 81.9) | 75.0 (65.4, 83.5) | **75.3 (59.4, 91.3)** |
| LLaVA | **85.4** | 67.6 | 25.9% | **86.3** | **85.9** | 60.4 |

| **Language** | | | | | | |
|---|---|---|---|---|---|---|
| Model | Overall | ES-AUC | Max Disp. | English | Spanish | Other |
| WResNet | 78.2 (74.9, 81.4) | 67.6 (59.4, 74.7) | **11.0%** | 76.3 (72.4, 80.0) | 87.3 (79.5, 93.5) | **83.0 (70.7, 91.9)** |
| ViT | 82.6 (79.9, 85.1) | 70.9 (61.4, 78.4) | 15.7% | 81.8 (78.8, 84.6) | 89.5 (85.8, 92.7) | 73.8 (56.7, 90.2) |
| EfficientNet | 78.7 (75.3, 81.8) | 69.4 (58.5, 76.3) | 13.1% | 78.4 (74.8, 81.9) | 79.2 (69.5, 87.5) | 66.1 (49.1, 81.6) |
| LLaVA | **85.4** | **71.3** | 18.8% | **84.5** | **93.4** | 74.6 |

**Table S6: Baseline model performance for advanced age-related macular degeneration (no fairness training).** AUCs and ES-AUC with 95% bootstrap CIs (2,000 resamples); Max Disp. = max between-subgroup AUC difference (point). LLaVA values are point estimates without CIs. Best per column **bold**.

**Sex**

| Model | Overall | ES-AUC | Max Disp. | Female | Male |
|---|---|---|---|---|---|
| WResNet | 83.0 (80.9, 85.0) | 77.3 (73.7, 81.0) | 7.3% | 80.0 (77.2, 82.8) | 87.3 (84.8, 89.6) |
| ViT | 84.7 (82.8, 86.5) | **81.8 (78.3, 85.1)** | **3.5%** | **83.3 (80.8, 85.8)** | 86.8 (84.3, 89.2) |
| EfficientNet | **86.3 (84.5, 88.1)** | 80.3 (76.8, 84.0) | 7.5% | **83.3 (80.6, 86.0)** | **90.8 (88.9, 92.5)** |
| LLaVA | 84.8 | 81.1 | 4.5% | 83.0 | 87.5 |

**Race**

| Model | Overall | ES-AUC | Max Disp. | Asian | Black | White | Other |
|---|---|---|---|---|---|---|---|
| WResNet | 83.0 (80.9, 85.0) | 51.5 (48.2, 72.5) | 55.9% | 42.0 (34.1, 49.8) | 97.9 (96.4, 99.1) | 83.4 (81.2, 85.5) | 78.2 (71.7, 84.3) |
| ViT | 84.7 (82.8, 86.5) | 60.5 (48.6, 77.7) | 35.6% | 62.2 (29.8, 93.7) | 97.8 (94.9, 100.0) | 85.0 (82.9, 86.9) | 80.5 (75.7, 85.2) |
| EfficientNet | **86.3 (84.5, 88.1)** | **63.1 (59.4, 76.1)** | **31.2%** | **68.4 (63.1, 73.7)** | **99.6 (98.6, 100.0)** | **86.5 (84.6, 88.3)** | **81.0 (74.5, 87.0)** |
| LLaVA | 84.8 | 61.2 | 32.8% | 65.7 | 98.5 | 85.0 | 79.3 |

**Ethnicity**

| Model | Overall | ES-AUC | Max Disp. | Non-Hisp | Hispanic | Other |
|---|---|---|---|---|---|---|
| WResNet | 83.0 (80.9, 85.0) | 71.9 (62.6, 78.6) | 15.0% | 82.6 (80.5, 84.6) | **93.0 (84.1, 98.4)** | 78.0 (62.4, 90.3) |
| ViT | 84.7 (82.8, 86.5) | **79.7 (70.6, 83.2)** | **5.5%** | 84.4 (82.4, 86.3) | 89.9 (81.5, 96.8) | **85.4 (75.1, 93.2)** |
| EfficientNet | **86.3 (84.5, 88.1)** | 77.6 (66.0, 83.2) | 11.0% | **86.5 (84.6, 88.3)** | 87.1 (72.2, 97.3) | 76.1 (65.1, 86.7) |
| LLaVA | 84.8 | 75.8 | 6.6% | 85.0 | 78.4 | 79.6 |

**Language**

| Model | Overall | ES-AUC | Max Disp. | English | Spanish | Other |
|---|---|---|---|---|---|---|
| WResNet | 83.0 (80.9, 85.0) | 70.7 (65.3, 75.6) | 13.2% | 82.2 (80.1, 84.3) | 95.4 (91.8, 98.5) | 87.2 (79.9, 93.6) |
| ViT | 84.7 (82.8, 86.5) | 74.9 (69.3, 81.3) | 8.4% | 84.1 (82.1, 86.0) | 92.5 (85.0, 98.4) | **89.3 (84.5, 93.7)** |
| EfficientNet | **86.3 (84.5, 88.1)** | 78.2 (71.9, 80.7) | 10.1% | **86.0 (84.0, 87.9)** | **96.0 (93.6, 98.2)** | 85.9 (78.1, 92.4) |
| LLaVA | 84.8 | **79.5** | **6.3%** | 84.5 | 82.0 | 88.3 |

**Table S7: Baseline model performance for glaucoma (no fairness training).** AUCs and ES-AUC with 95% bootstrap CIs (2,000 resamples); Max Disp. = max between-subgroup AUC difference (point). LLaVA values are point estimates without CIs. Best per column **bold**.

**Sex**

| Model | Overall | ES-AUC | Max Disp. | Female | Male |
|---|---|---|---|---|---|
| WResNet | 67.5 (65.5, 69.5) | 65.1 (62.0, 68.1) | 3.7% | 66.0 (63.3, 68.8) | 69.7 (66.8, 72.6) |
| ViT | **73.9 (71.9, 75.8)** | **72.0 (68.8, 74.8)** | 2.5% | **75.0 (72.4, 77.5)** | **72.5 (69.5, 75.4)** |
| EfficientNet | 67.7 (65.7, 69.6) | 67.4 (64.0, 68.9) | **0.3%** | 67.7 (65.0, 70.3) | 68.0 (65.0, 71.0) |

**Race**

| Model | Overall | ES-AUC | Max Disp. | Asian | Black | White | Other |
|---|---|---|---|---|---|---|---|
| WResNet | 67.5 (65.5, 69.5) | 50.2 (44.4, 55.5) | **32.4%** | 43.7 (31.5, 56.0) | 69.0 (59.4, 77.8) | 67.0 (64.8, 69.3) | 76.1 (70.4, 81.2) |
| ViT | **73.9 (71.9, 75.8)** | **54.9 (48.4, 61.1)** | 32.5% | **48.0 (34.6, 61.2)** | **72.0 (63.0, 80.6)** | **73.8 (71.6, 76.1)** | **80.5 (75.4, 85.1)** |
| EfficientNet | 67.7 (65.7, 69.6) | 47.7 (42.9, 51.8) | 39.2% | 39.5 (29.9, 49.8) | 65.5 (55.2, 75.1) | 67.1 (64.9, 69.4) | 78.7 (73.8, 83.3) |

**Ethnicity**

| Model | Overall | ES-AUC | Max Disp. | Non-Hisp | Hispanic | Other |
|---|---|---|---|---|---|---|
| WResNet | 67.5 (65.5, 69.5) | 54.3 (49.1, 59.7) | 23.6% | 66.8 (64.6, 68.9) | 82.6 (77.1, 87.3) | 59.0 (47.0, 70.1) |
| ViT | **73.9 (71.9, 75.8)** | **60.9 (55.6, 67.2)** | 20.9% | **73.5 (71.4, 75.6)** | **85.2 (79.3, 90.1)** | **64.3 (53.8, 75.0)** |
| EfficientNet | 67.7 (65.7, 69.6) | 56.1 (51.0, 60.4) | **19.9%** | 66.8 (64.7, 68.9) | 83.0 (77.2, 88.2) | 63.1 (53.8, 72.6) |

**Language**

| Model | Overall | ES-AUC | Max Disp. | English | Spanish | Other |
|---|---|---|---|---|---|---|
| WResNet | 67.5 (65.5, 69.5) | 55.0 (49.9, 57.7) | 22.4% | 66.4 (64.4, 68.4) | 88.8 (83.2, 93.7) | 67.0 (57.8, 75.8) |
| ViT | **73.9 (71.9, 75.8)** | **61.9 (57.2, 66.3)** | **16.3%** | **72.9 (70.9, 75.0)** | **89.2 (83.3, 94.4)** | **77.0 (69.9, 83.5)** |
| EfficientNet | 67.7 (65.7, 69.6) | 55.1 (50.4, 57.9) | 22.0% | 66.9 (64.8, 68.9) | 88.6 (83.5, 93.3) | 66.6 (58.9, 73.9) |

#### Demographics-only baseline, incremental value of the image, and paired fairness tests

These tables report the demographics-only baseline (Table S8), the incremental value of each image model over that baseline with its two-sided paired Student's $t$-test $p$-value (Table S9), and paired comparisons of each fairness method against the ViT baseline, reported as the change with its 95% bootstrap confidence interval (Table S10).

**Table S8:** Demographics-only baseline performance (logistic regression and gradient-boosted trees on age, race, and ethnicity). AUC with 95% bootstrap confidence intervals (100 patient-clustered resamples; $n$ = 20,380 test images).

| Disease | Baseline model | AUC (95% CI) |
|---|---|---|
| DR | Gradient-boosted trees | 74.2 (71.5–77.1) |
| DR | Logistic regression | 70.9 (67.2–75.7) |
| Advanced AMD | Gradient-boosted trees | 81.1 (79.7–82.6) |
| Advanced AMD | Logistic regression | 82.6 (81.0–84.0) |
| Glaucoma | Gradient-boosted trees | 55.0 (53.0–57.0) |
| Glaucoma | Logistic regression | 54.0 (52.4–56.4) |

**Table S9:** Incremental value of the image over demographics: difference between each image model's AUC and the better demographics-only baseline, with 95% bootstrap confidence intervals (100 patient-clustered resamples; $n = 20{,}380$ test images) and the two-sided paired Student's $t$-test $p$-value.

| Disease | Model | Image AUC (95% CI) | Demographics AUC (95% CI) | $\Delta$ (95% CI) | $p$ |
|---|---|---|---|---|---|
| DR | EfficientNet | 78.7 (76.0–81.3) | 74.2 (71.5–77.1) | +4.4 (+0.5, +8.1) | <0.05 |
| DR | ViT | 82.6 (80.3–85.3) | 74.2 (71.5–77.1) | +8.4 (+5.5, +11.8) | <0.001 |
| DR | WResNet | 78.2 (75.6–80.6) | 74.2 (71.5–77.1) | +4.0 (+0.4, +7.4) | <0.05 |
| Advanced AMD | EfficientNet | 86.3 (84.9–88.3) | 82.6 (81.0–84.0) | +3.8 (+2.2, +5.6) | <0.001 |
| Advanced AMD | ViT | 84.7 (82.8–86.5) | 82.6 (81.0–84.0) | +2.1 (+0.1, +3.8) | <0.05 |
| Advanced AMD | WResNet | 83.0 (81.3–85.3) | 82.6 (81.0–84.0) | +0.4 (−1.6, +2.3) | n.s. |
| Glaucoma | EfficientNet | 67.7 (65.9–69.4) | 55.0 (53.0–57.0) | +12.7 (+10.3, +14.8) | <0.001 |
| Glaucoma | ViT | 73.9 (72.3–75.7) | 55.0 (53.0–57.0) | +18.9 (+16.3, +21.2) | <0.001 |
| Glaucoma | WResNet | 67.5 (65.8–69.0) | 55.0 (53.0–57.0) | +12.4 (+10.3, +15.1) | <0.001 |

**Table S10:** Paired comparison of each fairness method against its baseline on the same test-set images (the ViT-based methods DRO, Adv, FSCL, FIS against the ViT baseline; FairLoRA against the LLaVA baseline). ΔES-AUC and ΔMax disparity are point-estimate changes (percentage points) in the equity-scaled AUC and in the maximum between-subgroup AUC difference, respectively; per-method ES-AUC with 95% bootstrap CIs is reported in Supplementary Tables S1–S2 (fairness) and S5–S6 (baseline). ΔOverall AUC is shown with its 95% bootstrap confidence interval. The sign indicates direction; a negative ΔMax disparity denotes reduced (improved) disparity.

| Disease | Attribute | Method | ΔES-AUC | ΔOverall AUC (95% CI) | ΔMax disparity |
|---|---|---|---|---|---|
| DR | Sex | DRO | −5.5 | −4.3 (−7.0, −1.8) | +1.7 |
| DR | Sex | Adv | +3.3 | +2.2 (+0.2, +3.5) | −1.4 |
| DR | Sex | FSCL | +1.1 | +0.4 (−1.6, +1.9) | −0.8 |
| DR | Sex | FIS | +4.0 | +3.1 (+0.8, +4.7) | −1.1 |
| DR | Sex | LLaVA+FairLoRA | −0.5 | – | −2.0 |
| DR | Race | DRO | +3.8 | −1.6 (−3.9, 0.0) | −12.4 |
| DR | Race | Adv | +18.7 | +0.8 (−1.9, +2.5) | −20.3 |
| DR | Race | FSCL | +16.4 | +1.4 (−0.9, +2.8) | −15.5 |
| DR | Race | FIS | +13.0 | +1.6 (−0.3, +2.9) | −12.8 |
| DR | Race | LLaVA+FairLoRA | +1.1 | – | −3.0 |
| DR | Ethnicity | DRO | +0.1 | +0.6 (−0.2, +1.5) | +0.8 |
| DR | Ethnicity | Adv | −0.2 | +1.4 (−0.9, +3.3) | +2.2 |
| DR | Ethnicity | FSCL | +1.9 | +0.7 (−1.4, +2.0) | −4.5 |
| DR | Ethnicity | FIS | +2.9 | +1.7 (−0.2, +3.5) | −3.0 |
| DR | Ethnicity | LLaVA+FairLoRA | +4.8 | – | −10.7 |
| DR | Language | DRO | −0.8 | −0.3 (−1.2, +0.5) | +0.7 |
| DR | Language | Adv | +0.8 | +1.1 (−1.4, +2.7) | −0.7 |
| DR | Language | FSCL | −1.4 | −0.1 (−2.3, +1.3) | +1.4 |
| DR | Language | FIS | +2.1 | +1.7 (−0.2, +3.4) | −1.5 |
| DR | Language | LLaVA+FairLoRA | +1.4 | – | −8.5 |
| AMD | Sex | DRO | −2.1 | −1.0 (−1.7, −0.3) | +1.5 |
| AMD | Sex | Adv | −1.2 | −0.3 (−1.0, +0.5) | +1.3 |
| AMD | Sex | FSCL | +0.1 | +0.8 (−0.4, +2.2) | +0.8 |
| AMD | Sex | FIS | −9.1 | −10.2 (−12.3, −7.9) | −1.1 |
| AMD | Sex | LLaVA+FairLoRA | −1.6 | – | +1.0 |
| AMD | Race | DRO | +7.0 | −1.7 (−3.4, −0.2) | −14.0 |
| AMD | Race | Adv | +0.9 | +0.3 (−0.3, +1.0) | −3.6 |
| AMD | Race | FSCL | −4.8 | +0.5 (−0.5, +1.8) | +7.1 |
| AMD | Race | FIS | −3.9 | −0.3 (−1.3, +0.8) | +9.3 |
| AMD | Race | LLaVA+FairLoRA | +0.4 | – | −3.5 |
| AMD | Ethnicity | DRO | −0.9 | −0.9 (−1.5, −0.2) | +0.7 |
| AMD | Ethnicity | Adv | −2.3 | −0.4 (−1.3, +0.3) | +3.3 |
| AMD | Ethnicity | FSCL | −9.4 | +1.2 (+0.3, +2.3) | +16.2 |
| AMD | Ethnicity | FIS | −1.9 | +0.2 (−0.9, +1.3) | +1.2 |
| AMD | Ethnicity | LLaVA+FairLoRA | −8.7 | – | +15.2 |
| AMD | Language | DRO | +2.8 | −0.2 (−0.8, +0.3) | −2.3 |
| AMD | Language | Adv | +0.8 | −0.1 (−1.0, +0.6) | 0.0 |
| AMD | Language | FSCL | +0.2 | +0.9 (0.0, +1.9) | +0.2 |
| AMD | Language | FIS | −1.0 | −0.4 (−1.4, +0.6) | +2.1 |
| AMD | Language | LLaVA+FairLoRA | −6.1 | – | +0.9 |

The complete per-subgroup fairness-training results for diabetic retinopathy and advanced age-related macular degeneration are reported in the consolidated tables above.

#### Subgroup case counts, age distribution, acquisition device and pupil dilation (Tables S11–S17)

Tables S11–S17 report disease-positive counts by self-reported race in the full cohort (Table S11), test-set image and positive-image counts by demographic subgroup (Table S12), the age distribution by disease status (Table S13), the distribution of images by acquisition-device group (Table S14), device-stratified AUCs of the baseline and fairness-trained models (Tables S15–S16), and AUCs stratified by pupil dilation status (Table S17).

**Table S11: Disease-positive counts by self-reported race in the full cohort (training and test sets combined).** Counts are given separately at the patient level (a patient is positive if any of their images carried a positive label) and at the image level; percentages are within-race prevalence. The glaucoma cohort is restricted to patients with visual-field data. Test-set image counts by subgroup are given in Table S12; patient-level counts by subgroup restricted to the test set were not available from the data export.

| Race | n | VTDR-positive, n (%) | Advanced AMD-positive, n (%) | Glaucoma cohort, n | Glaucoma-positive, n (%) |
|---|---|---|---|---|---|
| **Patients** | | | | | |
| Asian | 332 | 4 (1.2%) | 9 (2.7%) | 263 | 32 (12.2%) |
| Black | 407 | 22 (5.4%) | 4 (1.0%) | 289 | 34 (11.8%) |
| White | 7,916 | 53 (0.7%) | 287 (3.6%) | 6,655 | 605 (9.1%) |
| Other/Unknown | 1,223 | 36 (2.9%) | 27 (2.2%) | 1,054 | 67 (6.4%) |
| All | 9,878 | 115 (1.2%) | 327 (3.3%) | 8,261 | 738 (8.9%) |
| **Images** | | | | | |
| Race | n | VTDR-positive, n (%) | Advanced AMD-positive, n (%) | Glaucoma cohort, n | Glaucoma-positive, n (%) |
| Asian | 1,062 | 13 (1.2%) | 10 (0.9%) | 795 | 54 (6.8%) |
| Black | 1,429 | 70 (4.9%) | 5 (0.3%) | 986 | 57 (5.8%) |
| White | 33,422 | 133 (0.4%) | 668 (2.0%) | 27,489 | 1,194 (4.3%) |
| Other/Unknown | 4,774 | 99 (2.1%) | 54 (1.1%) | 4,052 | 117 (2.9%) |
| All | 40,687 | 315 (0.8%) | 737 (1.8%) | 33,322 | 1,422 (4.3%) |

**Table S12: Test-set image counts and positive-image counts by demographic subgroup.** The DR and advanced AMD test set comprised 20,380 images (193 VTDR-positive; 352 advanced AMD-positive); the glaucoma test set comprised 16,635 images (701 glaucoma-positive). Positive counts are images, not unique patients; because several images per patient were typically available, the number of independent positive patients within each subgroup is smaller than the number of positive images, and in the smallest strata (e.g., advanced AMD in Asian and Black patients) corresponds to at most one or two patients. Subgroup AUCs and ES-AUC values that depend on these strata should be regarded as exploratory.

| Attribute | Subgroup | Test images (DR/AMD) | VTDR-positive images | AMD-positive images | Test images (glaucoma) | Glaucoma-positive images |
|---|---|---|---|---|---|---|
| Sex | Female | 11,470 | 94 | 214 | 9,304 | 390 |
| | Male | 8,910 | 99 | 138 | 7,331 | 311 |
| Race | Asian | 525 | 7 | 2 | 432 | 27 |
| | Black | 727 | 45 | 2 | 505 | 34 |
| | White | 16,692 | 75 | 320 | 13,643 | 572 |
| | Other/Unknown | 2,436 | 66 | 28 | 2,055 | 68 |
| Ethnicity | Non-Hispanic | 18,532 | 155 | 334 | 15,139 | 636 |
| | Hispanic | 1,041 | 29 | 9 | 828 | 35 |
| | Unknown/Other | 807 | 9 | 9 | 668 | 30 |
| Primary language | English | 17,985 | 158 | 322 | 14,571 | 637 |
| | Spanish | 766 | 26 | 7 | 645 | 15 |
| | Other/Unknown | 1,629 | 9 | 23 | 1,419 | 49 |
| All | All | 20,380 | 193 | 352 | 16,635 | 701 |

**Table S13: Age distribution of the full cohort by disease status.** Ages in years. Patient-level rows use age at the patient's first study image; the image-level row uses age at image acquisition. Disease-positive groups may overlap. No ages were missing. Age bands by training/test split are given in Table 1 of the main text.

| Group | n | Mean (SD) | Median (IQR) | <18 | 18–39 | 40–64 | 65–79 | ≥80 |
|---|---|---|---|---|---|---|---|---|
| All patients (age at first image) | 9,878 | 59.7 (18.8) | 62.9 (48.9–73.1) | 306 (3.1%) | 1,283 (13.0%) | 3,795 (38.4%) | 3,282 (33.2%) | 1,212 (12.3%) |
| VTDR-positive patients | 115 | 58.9 (11.9) | 61.5 (51.4–66.3) | 0 (0.0%) | 10 (8.7%) | 67 (58.3%) | 35 (30.4%) | 3 (2.6%) |
| Advanced AMD-positive patients | 327 | 77.5 (9.6) | 78.5 (72.2–84.5) | 0 (0.0%) | 0 (0.0%) | 28 (8.6%) | 161 (49.2%) | 138 (42.2%) |
| Glaucoma-positive patients | 738 | 58.6 (19.4) | 61.3 (45.7–73.8) | 17 (2.3%) | 127 (17.2%) | 263 (35.6%) | 237 (32.1%) | 94 (12.7%) |
| All images (age at acquisition) | 40,687 | 61.7 (17.7) | 64.8 (52.2–74.1) | 835 (2.1%) | 4,347 (10.7%) | 15,390 (37.8%) | 14,632 (36.0%) | 5,483 (13.5%) |

#### Supplementary Note: performance by acquisition device

Topcon-labelled acquisitions accounted for 40,174 of 40,687 images (98·7%); the remaining images were labelled Zeiss (130), RetCam (17) or Optos (2), or carried no device label (364) (Table S14). Device groups were inferred from device labels embedded in the image file names and represent broad acquisition platforms rather than confirmed camera models. Within the Topcon group, test-set AUCs of the baseline ViT were 82·9% (95% CI 80·2–85·4) for vision-threatening DR, 85·5% (83·6–87·2) for advanced AMD and 73·9% (71·9–75·8) for glaucoma, compared with 82·6%, 84·7% and 73·9% across all devices (Table S15). The other device groups contained too few positive test images to support reliable estimates: the Zeiss group had one VTDR-positive and 11 AMD-positive test images (AMD AUC 36·1%, 95% CI 19·1–54·5, for ViT and 69·0%, 50·1–85·3, for EfficientNet), and the RetCam group contributed 17 images in total (three in the test set) with no positive labels. Fairness-trained ViT models behaved in the same way within the Topcon group (Table S16). Generalisability across devices is therefore not established.

**Table S14: Distribution of images by acquisition-device group, data split, and outcome class.** Device groups were inferred from device labels embedded in the image file names and represent broad acquisition platforms (Topcon: Maestro and Topcon colour retinal cameras; Zeiss: Clarus, Visucam and Visucam NM/FA; Optos: Silverstone; RetCam: Natus RetCam; Unspecified: no device label), not confirmed individual camera models. Counts are images. The glaucoma cohort is the subset with visual-field data.

| | **Training set (DR/AMD cohort)** | | | **Test set (DR/AMD cohort)** | | | **Training set (glaucoma)** | | **Test set (glaucoma)** | |
|---|---|---|---|---|---|---|---|---|---|---|
| Device group | Images | VTDR-positive | AMD-positive | Images | VTDR-positive | AMD-positive | Images | Glaucoma-positive | Images | Glaucoma-positive |
| Topcon | 20,045 | 109 | 383 | 20,129 | 192 | 341 | 16,468 | 719 | 16,419 | 693 |
| Zeiss | 80 | 9 | 2 | 50 | 1 | 11 | 68 | 2 | 34 | 1 |
| Optos | 2 | 1 | 0 | 0 | 0 | 0 | 1 | 0 | 0 | 0 |
| RetCam | 14 | 0 | 0 | 3 | 0 | 0 | 14 | 0 | 3 | 0 |
| Unspecified | 166 | 3 | 0 | 198 | 0 | 0 | 136 | 0 | 179 | 7 |
| All | 20,307 | 122 | 385 | 20,380 | 193 | 352 | 16,687 | 721 | 16,635 | 701 |

**Table S15: Device-stratified test-set AUC (95% bootstrap CI) of the baseline models.** Device groups as in Table S14. Cells marked n.e. (not evaluable) contain fewer than two positive images (Zeiss: one VTDR-positive and one glaucoma-positive image; Unspecified: no VTDR- or AMD-positive images) and therefore cannot support an AUC; RetCam (3 test images, no positives) and Optos (no test images) are omitted. Image counts (positive images) per device group are shown in the sub-header for each task. Confidence intervals in this table and in Table S16 were computed in a separate stratified bootstrap run and may differ marginally from those in Tables S1–S7.

| Task | Model | Topcon | Zeiss | Unspecified | All devices |
|---|---|---|---|---|---|
| *VTDR test images (positive): Topcon 20,129 (192); Zeiss 50 (1); Unspecified 198 (0); all devices 20,380 (193)* | | | | | |
| VTDR | WResNet | 78.6 (75.3, 81.9) | n.e. | n.e. | 78.2 (74.9, 81.6) |
| | ViT | 82.9 (80.2, 85.4) | n.e. | n.e. | 82.6 (80.1, 85.1) |
| | EfficientNet | 79.0 (75.7, 82.1) | n.e. | n.e. | 78.7 (75.5, 81.8) |
| *Advanced AMD test images (positive): Topcon 20,129 (341); Zeiss 50 (11); Unspecified 198 (0); all devices 20,380 (352)* | | | | | |
| Advanced AMD | WResNet | 84.0 (82.1, 85.9) | 39.2 (22.9, 56.5) | n.e. | 83.0 (81.0, 84.9) |
| | ViT | 85.5 (83.6, 87.2) | 36.1 (19.1, 54.5) | n.e. | 84.7 (82.9, 86.5) |
| | EfficientNet | 87.1 (85.4, 88.8) | 69.0 (50.1, 85.3) | n.e. | 86.3 (84.5, 88.1) |
| *Glaucoma test images (positive): Topcon 16,419 (693); Zeiss 34 (1); Unspecified 179 (7); all devices 16,635 (701)* | | | | | |
| Glaucoma | WResNet | 67.7 (65.6, 69.7) | n.e. | 37.4 (15.2, 62.1) | 67.5 (65.5, 69.4) |
| | ViT | 73.9 (71.9, 75.8) | n.e. | 70.8 (37.8, 97.4) | 73.9 (71.9, 75.9) |
| | EfficientNet | 68.0 (66.0, 69.9) | n.e. | 37.4 (20.2, 55.0) | 67.7 (65.6, 69.6) |

**Table S16: Device-stratified test-set AUC (95% bootstrap CI) of the fairness-trained ViT models.** The ViT baseline is shown in Table S15. Test images (positive images) per device group: VTDR – Topcon 20,129 (192), Zeiss 50 (1), all devices 20,380 (193); advanced AMD – Topcon 20,129 (341), Zeiss 50 (11), all devices 20,380 (352). n.e. = not evaluable (fewer than two positive images). The Unspecified group (198 test images) contained no VTDR- or AMD-positive images and is omitted.

| Task | Attribute | Method | Topcon | Zeiss | All devices |
|---|---|---|---|---|---|
| VTDR | Sex | ViT + DRO | 78.7 (75.5, 81.7) | n.e. | 78.4 (75.4, 81.4) |
| | | ViT + Adv | 85.3 (82.7, 87.8) | n.e. | 84.8 (82.2, 87.3) |
| | | ViT + FSCL | 83.4 (80.8, 86.0) | n.e. | 83.0 (80.3, 85.6) |
| | | ViT + FIS | 86.0 (83.4, 88.4) | n.e. | 85.7 (83.0, 88.3) |
| VTDR | Race | ViT + DRO | 81.4 (78.5, 84.1) | n.e. | 81.0 (78.3, 83.9) |
| | | ViT + Adv | 83.5 (80.4, 86.3) | n.e. | 83.4 (80.3, 86.1) |
| | | ViT + FSCL | 84.3 (81.6, 86.7) | n.e. | 84.1 (81.4, 86.6) |
| | | ViT + FIS | 84.5 (81.7, 87.2) | n.e. | 84.2 (81.3, 86.7) |
| VTDR | Ethnicity | ViT + DRO | 83.5 (81.0, 86.0) | n.e. | 83.2 (80.5, 85.9) |
| | | ViT + Adv | 84.4 (81.4, 87.1) | n.e. | 84.0 (81.1, 87.0) |
| | | ViT + FSCL | 83.6 (80.8, 86.1) | n.e. | 83.3 (80.7, 85.8) |
| | | ViT + FIS | 84.6 (81.8, 87.2) | n.e. | 84.3 (81.6, 87.0) |
| VTDR | Language | ViT + DRO | 82.6 (79.8, 85.2) | n.e. | 82.3 (79.7, 84.8) |
| | | ViT + Adv | 84.1 (81.4, 86.9) | n.e. | 83.7 (80.9, 86.3) |
| | | ViT + FSCL | 82.7 (80.0, 85.4) | n.e. | 82.5 (79.7, 85.1) |
| | | ViT + FIS | 84.6 (82.0, 87.1) | n.e. | 84.3 (81.7, 86.7) |
| Advanced AMD | Sex | ViT + DRO | 84.6 (82.8, 86.4) | 38.0 (17.3, 62.7) | 83.7 (81.8, 85.6) |
| | | ViT + Adv | 85.3 (83.5, 86.9) | 31.2 (11.4, 51.2) | 84.4 (82.5, 86.2) |
| | | ViT + FSCL | 85.9 (83.9, 87.7) | 50.1 (26.6, 74.5) | 85.4 (83.5, 87.3) |
| | | ViT + FIS | 75.7 (73.2, 78.1) | 36.6 (15.9, 58.3) | 74.4 (71.8, 77.0) |
| Advanced AMD | Race | ViT + DRO | 83.9 (82.0, 85.8) | 38.0 (14.0, 62.7) | 83.0 (80.9, 85.0) |
| | | ViT + Adv | 85.9 (84.0, 87.7) | 35.4 (17.1, 55.7) | 85.0 (83.1, 86.9) |
| | | ViT + FSCL | 86.1 (84.2, 87.8) | 35.9 (15.8, 58.1) | 85.2 (83.2, 87.1) |
| | | ViT + FIS | 85.4 (83.7, 87.2) | 26.8 (10.0, 45.8) | 84.4 (82.6, 86.2) |
| Advanced AMD | Ethnicity | ViT + DRO | 84.7 (82.8, 86.4) | 44.1 (25.2, 64.0) | 83.8 (81.8, 85.6) |
| | | ViT + Adv | 85.3 (83.2, 87.0) | 29.1 (10.6, 50.4) | 84.3 (82.3, 86.1) |
| | | ViT + FSCL | 86.8 (85.1, 88.5) | 35.2 (16.4, 57.1) | 85.9 (84.1, 87.6) |
| | | ViT + FIS | 85.6 (83.6, 87.3) | 35.0 (16.3, 55.1) | 84.9 (83.1, 86.7) |
| Advanced AMD | Language | ViT + DRO | 85.4 (83.6, 87.2) | 36.8 (17.6, 58.5) | 84.5 (82.6, 86.2) |
| | | ViT + Adv | 85.5 (83.5, 87.3) | 29.8 (11.0, 50.9) | 84.5 (82.5, 86.3) |
| | | ViT + FSCL | 86.6 (84.7, 88.4) | 34.7 (15.2, 58.3) | 85.6 (83.8, 87.3) |
| | | ViT + FIS | 85.0 (83.2, 86.8) | 40.6 (20.6, 61.8) | 84.3 (82.4, 86.1) |

**Table S17: Test-set AUC (95% bootstrap CI) stratified by pupil dilation status at image acquisition.** Dilation status was assigned during manual image review (dilated versus undilated external photographs). Models were not retrained; the trained models were evaluated separately on the dilated and undilated subsets of the test set. Test images (positive images): VTDR/advanced AMD – all 20,380 (193 VTDR; 352 AMD), dilated 19,523 (172; 326), undilated 857 (21; 26); glaucoma – all 16,635 (701), dilated 15,981 (677), undilated 654 (24). The undilated subset is small and its confidence intervals are correspondingly wide.

| Task | Attribute | Model | All images | Dilated | Undilated |
|---|---|---|---|---|---|
| VTDR | Baseline | WResNet | 78.2 (74.9, 81.4) | 78.0 (74.3, 81.3) | 73.1 (62.4, 82.8) |
| | | ViT | 82.6 (79.9, 85.1) | 83.3 (80.4, 85.8) | 70.8 (58.8, 82.2) |
| | | EfficientNet | 78.7 (75.3, 81.8) | 79.3 (76.0, 82.4) | 61.1 (46.5, 75.4) |
| Advanced AMD | Baseline | WResNet | 83.0 (80.9, 85.0) | 84.3 (82.3, 86.1) | 66.4 (57.2, 75.9) |
| | | ViT | 84.7 (82.8, 86.5) | 85.7 (83.8, 87.4) | 74.0 (65.6, 82.2) |
| | | EfficientNet | 86.3 (84.5, 88.1) | 87.2 (85.3, 89.0) | 76.2 (66.5, 85.1) |
| Glaucoma | Baseline | WResNet | 67.5 (65.5, 69.5) | 67.4 (65.3, 69.4) | 68.0 (59.4, 76.4) |
| | | ViT | 73.9 (71.9, 75.8) | 74.0 (71.9, 76.0) | 72.0 (62.2, 80.5) |
| | | EfficientNet | 67.7 (65.7, 69.6) | 67.8 (65.8, 69.8) | 65.8 (54.5, 76.6) |
| VTDR | Sex | ViT + DRO | 78.4 (75.2, 81.2) | 77.6 (74.4, 80.9) | 79.1 (70.1, 87.3) |
| | | ViT + Adv | 84.8 (82.2, 87.2) | 84.7 (81.9, 87.4) | 79.2 (71.0, 86.6) |
| | | ViT + FSCL | 83.0 (80.3, 85.5) | 83.0 (80.1, 85.8) | 76.8 (67.5, 85.5) |
| | | ViT + FIS | 85.7 (83.1, 88.1) | 85.6 (82.7, 88.3) | 81.3 (73.3, 88.4) |
| VTDR | Race | ViT + DRO | 81.0 (78.1, 83.8) | 81.1 (77.9, 84.0) | 67.7 (57.9, 76.5) |
| | | ViT + Adv | 83.4 (80.3, 86.3) | 82.9 (79.5, 86.2) | 79.9 (72.0, 87.4) |
| | | ViT + FSCL | 84.1 (81.3, 86.5) | 84.2 (81.2, 87.2) | 74.6 (64.6, 83.4) |
| | | ViT + FIS | 84.2 (81.3, 86.7) | 84.0 (81.0, 86.8) | 79.7 (70.5, 87.7) |
| VTDR | Ethnicity | ViT + DRO | 83.2 (80.4, 85.7) | 83.8 (80.9, 86.5) | 71.0 (59.9, 81.6) |
| | | ViT + Adv | 84.0 (81.1, 86.7) | 84.0 (81.0, 87.0) | 79.8 (70.9, 87.5) |
| | | ViT + FSCL | 83.3 (80.6, 85.8) | 83.4 (80.5, 86.3) | 75.5 (66.3, 83.9) |
| | | ViT + FIS | 84.3 (81.5, 86.9) | 84.1 (81.2, 87.0) | 80.6 (70.2, 89.0) |
| VTDR | Language | ViT + DRO | 82.3 (79.6, 84.8) | 82.9 (80.0, 85.6) | 71.7 (60.0, 82.7) |
| | | ViT + Adv | 83.7 (80.7, 86.4) | 83.6 (80.6, 86.7) | 80.1 (71.9, 87.6) |
| | | ViT + FSCL | 82.5 (79.6, 85.1) | 82.7 (79.6, 85.6) | 77.8 (68.5, 86.7) |
| | | ViT + FIS | 84.3 (81.5, 86.8) | 84.6 (81.7, 87.3) | 73.9 (61.7, 85.0) |
| Advanced AMD | Sex | ViT + DRO | 83.7 (81.8, 85.5) | 84.9 (83.1, 86.7) | 74.8 (67.0, 82.7) |
| | | ViT + Adv | 84.4 (82.5, 86.3) | 85.6 (83.8, 87.3) | 73.5 (65.0, 82.3) |
| | | ViT + FSCL | 85.4 (83.6, 87.2) | 86.3 (84.4, 88.1) | 78.7 (70.3, 85.9) |
| | | ViT + FIS | 74.4 (71.9, 77.1) | 75.0 (72.3, 77.6) | 66.7 (57.0, 76.5) |
| Advanced AMD | Race | ViT + DRO | 83.0 (80.9, 85.0) | 84.4 (82.4, 86.3) | 68.8 (59.2, 77.8) |
| | | ViT + Adv | 85.0 (83.1, 86.9) | 86.0 (84.1, 87.7) | 73.5 (64.2, 82.7) |
| | | ViT + FSCL | 85.2 (83.2, 87.0) | 86.1 (84.2, 87.9) | 76.2 (66.9, 85.2) |
| | | ViT + FIS | 84.4 (82.5, 86.2) | 85.7 (83.9, 87.4) | 73.3 (64.9, 81.6) |
| Advanced AMD | Ethnicity | ViT + DRO | 83.8 (81.8, 85.7) | 84.9 (83.0, 86.8) | 71.3 (62.1, 80.5) |
| | | ViT + Adv | 84.3 (82.2, 86.2) | 85.5 (83.6, 87.4) | 71.9 (63.0, 80.8) |
| | | ViT + FSCL | 85.9 (84.1, 87.6) | 86.9 (85.1, 88.6) | 75.9 (67.8, 84.0) |
| | | ViT + FIS | 84.9 (83.1, 86.7) | 85.6 (83.8, 87.4) | 74.0 (64.9, 82.5) |
| Advanced AMD | Language | ViT + DRO | 84.5 (82.6, 86.4) | 85.5 (83.7, 87.3) | 71.9 (62.1, 81.2) |
| | | ViT + Adv | 84.5 (82.5, 86.5) | 85.9 (84.1, 87.7) | 72.6 (63.9, 81.1) |
| | | ViT + FSCL | 85.6 (83.8, 87.4) | 86.5 (84.8, 88.3) | 77.1 (69.2, 85.1) |
| | | ViT + FIS | 84.3 (82.4, 86.1) | 85.2 (83.3, 87.0) | 76.1 (67.5, 83.9) |
